# Population-scale disease-associated tandem repeat analysis reveals locus and ancestry-specific insights

**DOI:** 10.1101/2025.10.11.25337795

**Authors:** Indhu-Shree Rajan-Babu, Readman Chiu, Ben Weisburd, Iris Caglayan, Inanc Birol, Jan M. Friedman

**Author notes:** **Corresponding author:** Indhu-Shree Rajan-Babu, Ph.D. Department of Medical Genetics The University of British Columbia 4500 Oak Street, Vancouver, BC V6H 3N1, Canada |.

## Abstract

Tandem repeat (TR) expansions underlie many monogenic disorders, with variable length and sequence influencing pathogenicity, disease penetrance, severity, and onset. Accurate genotype-phenotype correlation and disease prevalence estimation require molecular characterization beyond repeat length. Here we present a population-scale analysis of 66 disease-associated TR loci using long-read assemblies from 2,526 diverse haplotypes. Integrating repeat length, motif composition, local ancestry, linkage disequilibrium, and phylogenetic analyses, we reveal extensive locus-, population-, and allele-specific variation shaping disease risk. Up to 16% of individuals have one or more locus with repeat numbers above established pathogenic thresholds. Many of these expansions contain interrupting motifs or novel sequence structures attenuating pathogenicity, highlighting the need to refine screening and diagnostic criteria beyond repeat length alone. Our results demonstrate that polymorphic enlarged alleles with incomplete or no clinical penetrance may occur at some disease-associated TR loci. Ancestry-resolved analyses uncover population-specific TR architectures contributing to epidemiological disparities in repeat expansion disorders. Phylogenetic analyses identify conserved ancestral alleles and loci with recent instability and mutation rates influenced by selective pressures. We also describe variable linkage disequilibrium patterns and recombination signatures around specific disease-associated TR loci. Our findings emphasize integrating sequence, ancestry, and evolutionary context to understand disease-associated TR loci’s complex landscape.

## Introduction

Disorders caused by tandem repeat (TR) expansions that exceed locus-specific thresholds at approximately 70 loci^1^ affect roughly 1 in 3,000 individuals worldwide^2^. While most repeat expansion (RE) disorders occur across diverse ancestries, some are population-specific or notably prevalent in particular groups^3^.

Disease-associated REs (“full mutations”) exhibit substantial heterogeneity in length and sequence composition, including interruptions and non-canonical motifs that vary by locus and population background and can critically influence disease risk, onset, penetrance, and inheritance^4^. At certain loci, pathogenicity is also associated with RE-triggered methylation-mediated loss of gene function^5^.

Haplotype structures surrounding TR loci further influence repeat stability and the feasibility of imputation or genetic screening^6,7^. Importantly, these genomic contexts vary considerably across populations due to recombination, selection, genetic drift, and admixture^8^. Such processes can result in local ancestry at a TR locus that differs from an individual’s genome-wide ancestry, further complicating disease allele distribution and definition.

Recent analysis of a subset of TR loci in short-read sequencing data reported a ∼10× higher RE carrier frequency and a 2–3× higher disorder prevalence than previously recognized, and identified numerous premutation carriers at risk of having affected children^3^. However, short-read sequencing cannot reliably size REs that are longer than sequencing read lengths (approximately 150–350 base pairs) or detect changes in methylation and sequence composition—the latter known to modify disease at approximately a third of loci^4^. Accurate estimation of pathogenic RE prevalence and disease risk requires comprehensive assessment of TRs and their genomic context.

Long-read sequencing (LRS) produces reads tens of kilobases in length, enabling direct, base-level characterization of TR length, sequence composition, interruptions, and associated methylation changes in a single assay—offering unparalleled capabilities for accurate RE assessment^9,10^. Previous LRS-based studies have cataloged population-level TR variation^9,11–15^; however, some are restricted to a specific population, limiting insights into ancestrally diverse cohorts, and/or often lack comprehensive characterization of repeat heterogeneity—including population-specific alleles, motif structures, genomic context, or local ancestry—at clinically relevant loci. Thus, a detailed, population- and locus-specific analysis incorporating genomic context is urgently needed to enhance disease risk modeling and mechanistic understanding of TR stability.

To address these gaps, we present a comprehensive, ancestry-informed analysis of 66 known disease-associated TR loci using 2,526 phased haplotypes from LRS datasets generated by the Human Pangenome Reference Consortium (HPRC)^16^, the 1000 Genomes Project ONT (1kGP-ONT) Sequencing Consortium^12^, the Human Genome Structural Variation Consortium (Phase 2; HGSVC2)^17^, and the 1kGP individuals sequenced by Noyvert *et al*^18^. Integrating TR length, sequence motif composition, linkage disequilibrium (LD) patterns, recombination, and local ancestry, we delineate locus- and population-specific TR architectures, assess the clinical significance of expanded alleles, and compare human TR sequences to primate orthologs. Our analysis reveals ancestral and recently unstable loci, identifies population-specific alleles underlying epidemiological disparities, and highlights the evolutionary dynamics shaping TR variation. This work establishes a foundational framework for interpreting RE pathogenicity, and evolution by mapping the complex locus-, population-, and allele-specific trajectories of disease-associated TRs in an integrated genomic context, advancing genetic screening, clinical interpretation, and understanding of epidemiological disparities in the distribution of RE disorders, and enhancing our understanding of genome biology.

## Results

### Locus- and Population-Specific TR Diversity

We genotyped 66 disease-associated TR loci (Fig. 1a) in 1263 healthy individuals from two cohorts, stratified by superpopulation (Africans AFR; Admixed Americans AMR; East Asians EAS; Europeans EUR; South Asians SAS) to assess population-specific variation in TR lengths (Fig. 1b; Supplementary Fig. 1). TR alleles were classified as normal, intermediate, premutation, reduced-penetrance, full mutation, or “unknown” if outside defined size-based allelic categories. Cohort 1 (272 samples from HPRC, HGSVC2, and 1kGP-ONT) exhibited unimodal repeat-length distributions at several loci (e.g., *ARX-1*, *COMP*) (Supplementary Fig. 1), whereas the larger Cohort 2 (991 1kGP samples from Noyvert *et al*.) revealed greater allelic diversity (Fig. 1b). Inter-population differences in repeat-length distributions were significant at 34% of pairwise genotype comparisons in Cohort 2 and 12% in Cohort 1, with 58–66% of this variability attributable to AFR individuals (Supplementary Tables 1,2). Comparisons between cohorts revealed significant differences at eleven loci (Supplementary Table 3).

**Fig. 1.**
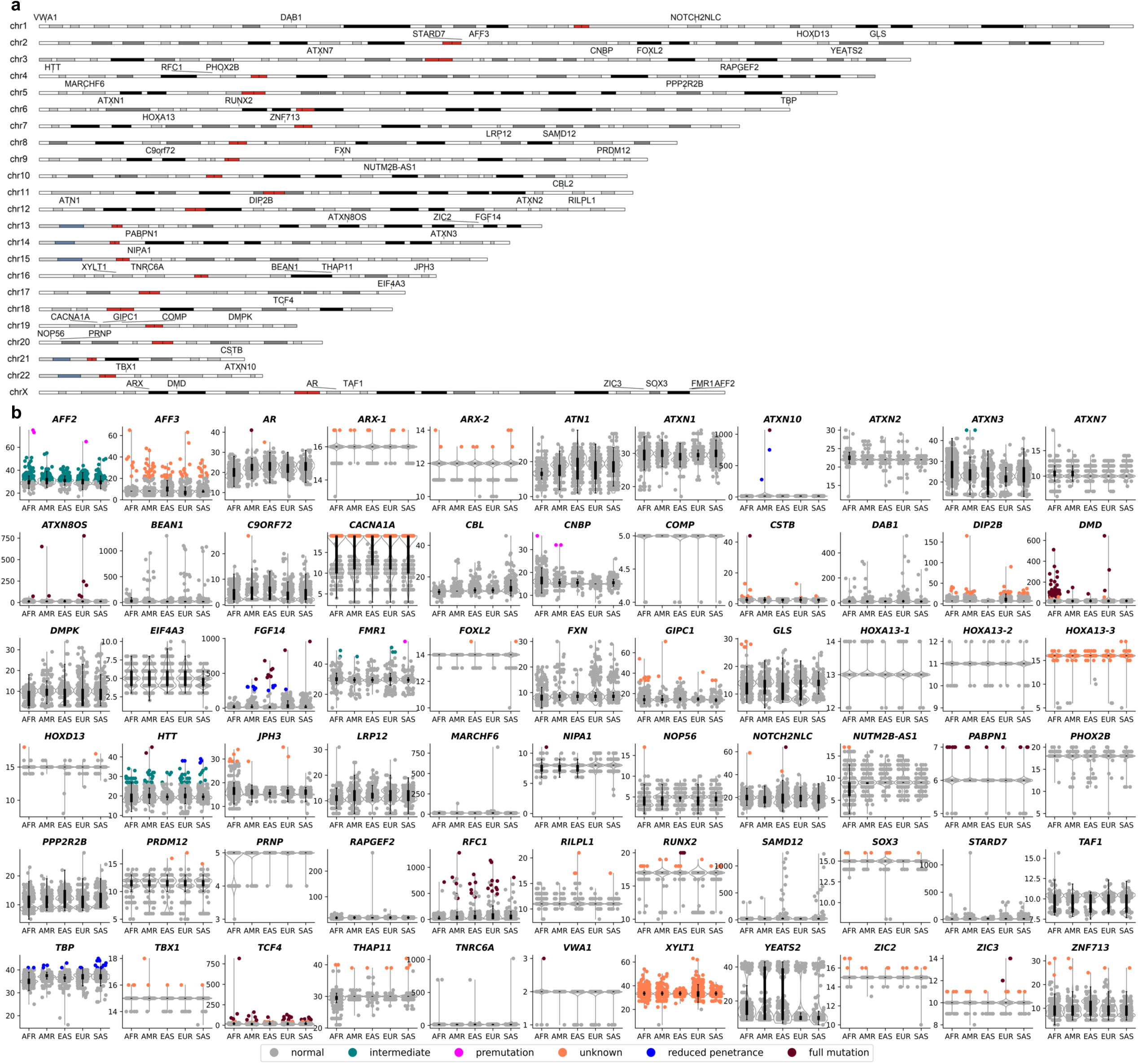
Chromosome-scale annotations of known disease-associated TR loci and allelic classifications and repeat length distributions in Cohort 2 (a,b). Genomic locations of 66 known disease-associated TR loci are shown across chromosomes 1-22 and X (**a**). Disease-associated TR loci are identified on all chromosomes except Y. Chromosome X contains the highest number (eight) of known disease-associated TRs, followed by chromosome 16 with five. Chromosomes 10, 11, 15, 17, 18, and 21 each harbor a single disease-associated TR each, while the remaining chromosomes contain two to four loci each. Raincloud plots show repeat length distributions across 66 known disease-associated TR loci, derived from *de novo* haplotype-resolved genome assemblies (**b**). The *y*-axis represents the estimated number of repeat copies. Genotypes are stratified by assigned ancestry or superpopulation (AFR, AMR, EUR, EAS, SAS) of the 991 individuals from Cohort 2. Scatter plot colors indicate the allelic class of each genotyped TR allele: normal, intermediate, premutation, unknown, reduced penetrance, or full-mutation.

Population-specific TR allele frequencies were evident at several loci. For example, the stable *DIP2B* 7-repeat allele was nearly fixed in EAS (98%), compared with SAS (82%), AFR and AMR (77% each), and was least frequent in EUR (58%), the only population where the RE-associated intellectual developmental disorder has been reported^19^. The overall repeat copy distribution in EAS also differed significantly from those in other populations (Fig. 2a). At *FGF14*, the repeat copy distribution differed significantly between all populations (Fig. 2b). In EAS, the normal allele distribution was tightly clustered (∼5–50 repeats), remaining below the 200–249 range associated with increased instability and intergeneration expansion to pathogenic sizes (>250)^20^, consistent with the lower prevalence of *FGF14*-associated ataxia in EAS^20,21^.

**Fig. 2.**
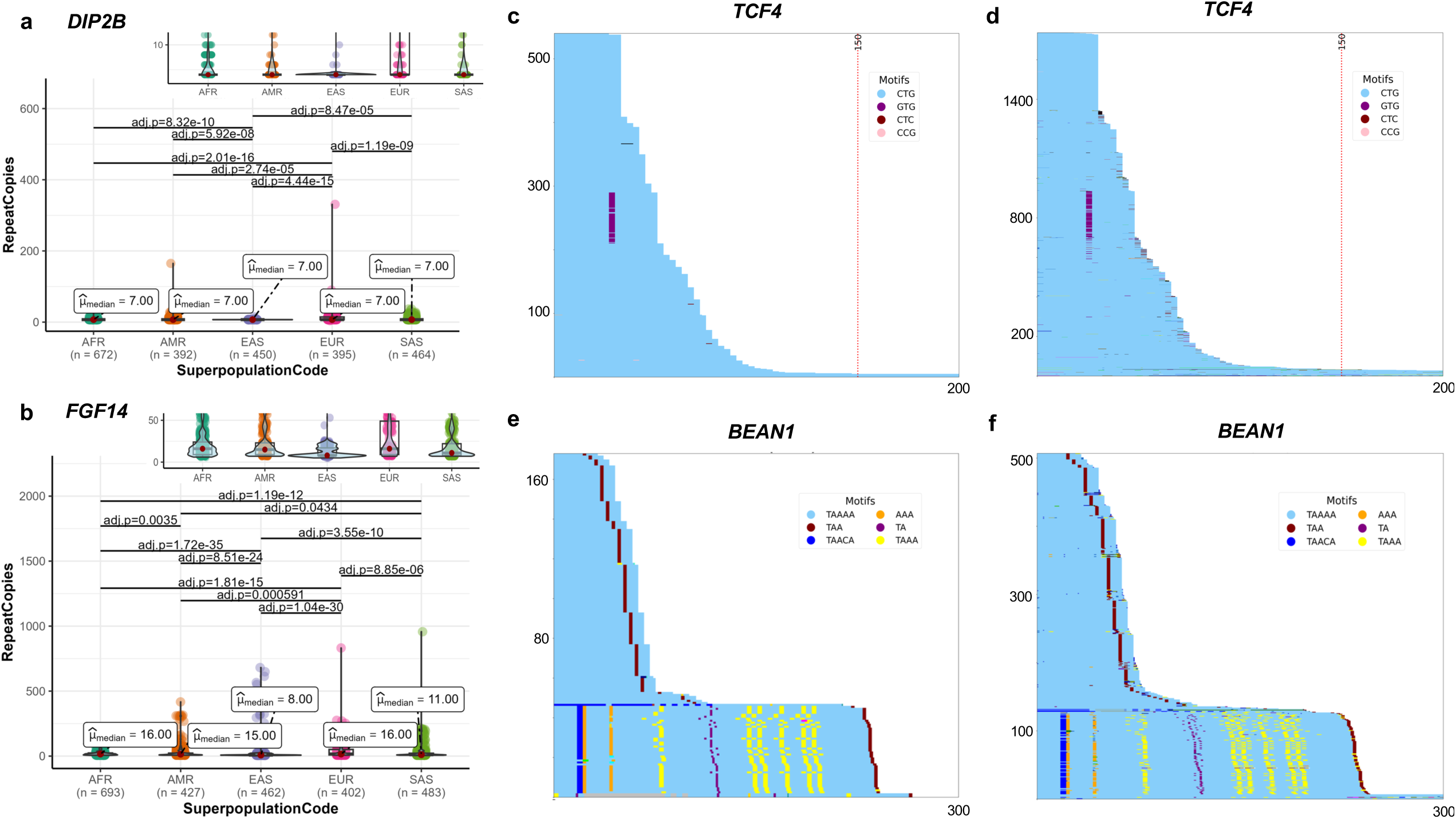
Repeat length distributions of *DIP2B* and *FGF14*, and sequence compositions of *TCF4* and *BEAN1* TRs in Cohorts 1 and 2 (a–f). Annotated violin–boxplots with raw data overlay show the distribution of repeat copies at *DIP2B* (**a**) and *FGF14* (**b**). The *y*-axis shows the estimated number of repeat copies, stratified by assigned ancestry or superpopulation (AFR, AMR, EUR, EAS, or SAS). Group medians are shown for each category. Statistically significant pairwise differences between superpopulations, as assessed by the Kolmogorov–Smirnov test with Bonferroni correction, are indicated above the corresponding group comparisons. Inset panels depict the zoomed-in distribution of normal alleles. Sequence compositions of *TCF4* (**c,d**) and *BEAN1* (**e,f**) loci are shown for Cohorts 1 (left) and 2 (right). The *TCF4* data include sequences from individuals of all superpopulations, while the *BEAN1* data include sequences only from individuals of AFR population. The canonical motif—CTG in *TCF4* and TAAAA in *BEAN1*—are shown in light sky blue, with identified non-canonical motifs and their colors indicated in the legend box of each panel. The *x*-axis shows repeat size in base pairs, and the *y*-axis shows the number of alleles characterized. A vertical red dotted line in the *TCF4* panels marks the reported threshold for the lower bound of full-mutation allele (∼50 CTG repeats). Vertical repeat threshold for *BEAN1* is not shown because the panel is zoomed in to highlight the composition of normal alleles. Only top recurring non-canonical motifs are shown in the figure legends.

Sequence composition analysis identified population-, locus-, and allele-specific TR structures. At *TCF4*, GTG interruption was highly enriched among alleles with 16–17 repeats (Fig. 2c,d), while at *BEAN1*, most alleles with 52–57 repeats in AFR individuals displayed a distinctive sequence of motifs: TAACA, two (A)_2-4_ monomers, TAAA, TA, five TAAAs, and TAA, alternating with canonical (TTTTA)_n_ (Fig. 2e,f), revealing population- and length-specific TR configurations.

At *ATXN2*, two CAA interruptions were found in 64% of normal (≤22 CAG) alleles but only in 46% of longer alleles, which often carried just one CAA interruption and featured longer uninterrupted 5’ (CAG)_n_ tracts (40%) (Fig. 3a), patterns that may influence repeat stability^22^.

**Fig. 3.**
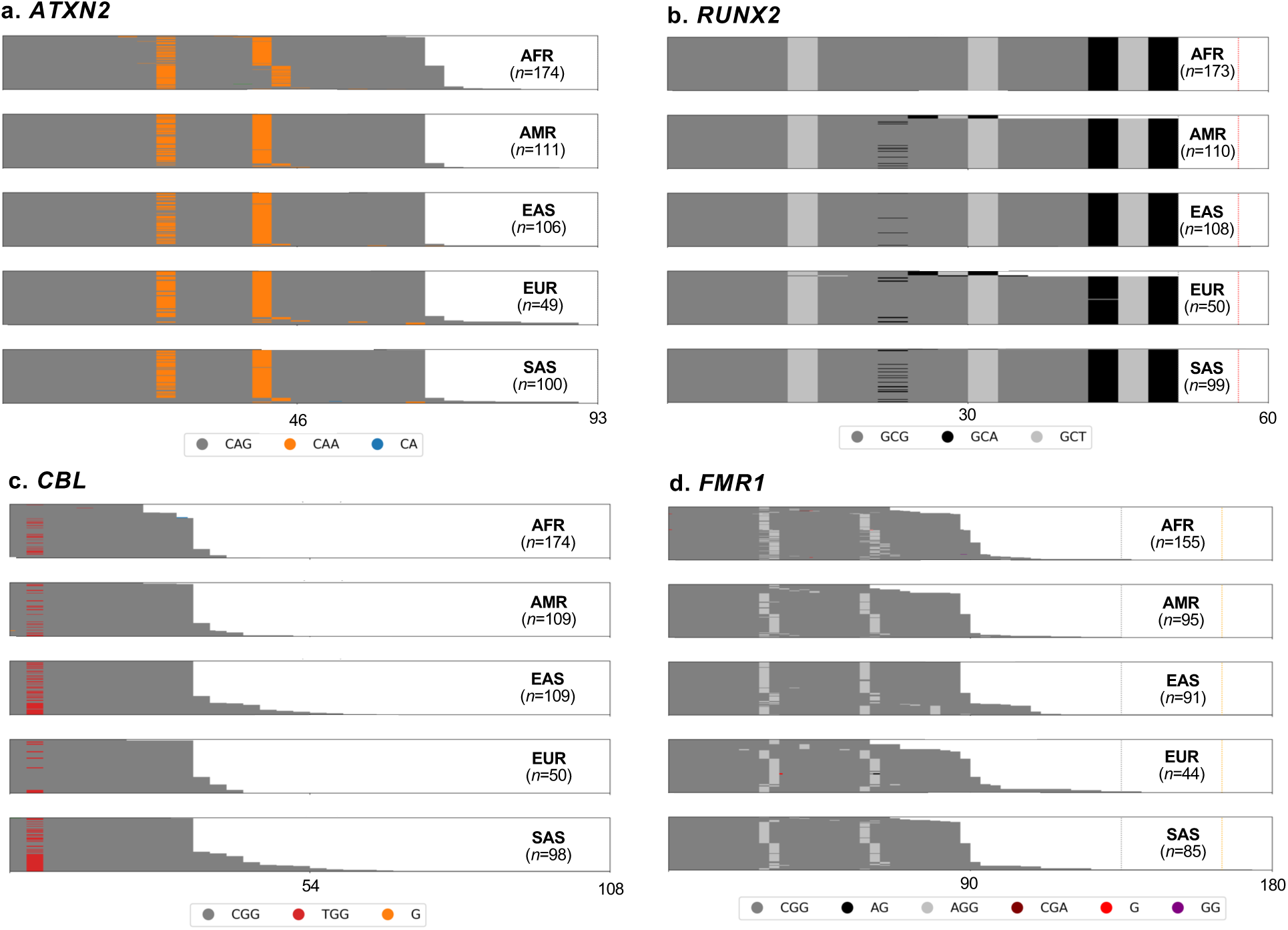
Sequence compositions of *ATXN2*, *CBL*, *FMR1*, and *RUNX2* TR loci stratified by superpopulation (a–d). The number of alleles characterized for each superpopulation or assigned ancestry (AFR, AMR, EAS, EUR, SAS) at each locus is indicated in the top right of each panel. Canonical motifs— CAG in *ATXN2* (**a**), GCG in *RUNX2* (**b**), and CGG in *CBL* (**c**) and *FMR1* (**d**)—are shown in dark gray. Identified non-canonical motifs and their corresponding colors are indicated in the legend below the panels. The *x*-axis denotes repeat size in base pairs. Vertical dotted lines in *RUNX2* (**b**) and *FMR1* (**d**) panels indicate reported repeat size thresholds: gray for the upper bounds of normal, orange for the lower bounds of premutation alleles, and red for full-mutation alleles. *ATXN2* (**a**) and *CBL* (**c**) panels do not include vertical threshold lines because they are zoomed in to highlight the composition of normal alleles. For clarity, only data from Cohort 1 is presented in this figure; analyses including both cohorts are reported in the main text.

Notably, the AFR population had a higher frequency (25%) of the 23-repeat allele with two CAA interruptions compared to ≤4% in other populations. At *RUNX2*, the most common structure was (GCG)_4_-GCT-(GCG)_5_-GCT-(GCG)_3_-GCA-GCT-GCA, whereas an alternate structure with (GCG)_2_-GCA-(GCG)_2_ in place of (GCG)_5_ was more prevalent in SAS (16.4%) compared to AFR (0.7%), EAS (3.3%), AMR (7.5%), and EUR (9.4%) (Fig. 3b), reflecting population-specific variation in sequence configuration. At *CBL*, longer alleles (>11 repeats) more often carried a TGG interruption in comparison to shorter alleles (AFR: 41% vs. 18%; AMR: 31% vs. 16%; EAS: 69% vs. 39%; EUR: 32% vs. 28%; SAS: 74% vs. 35%) (Fig. 3c), suggesting a consistent length-dependent sequence structure at this locus. At *FMR1*, alleles with >30 repeats were most frequent in AFR (35%) and least frequent in AMR (13%). In EAS, 44% of these alleles carried three AGG interruptions (notably higher than SAS (14%), AMR (7%), EUR and AFR (3% each)) predominantly in (CGG)_9_-AGG-(CGG)_9_-AGG-(CGG)_6_-AGG-(CGG)_9_ configuration, which was largely restricted to EAS and SAS (Fig. 3d), consistent with known regional patterns of *FMR1* AGG interruptions^23^. See Additional Data Figs. 1 and 2 for sequence composition plots of all loci in both cohorts.

Across loci, AFR individuals exhibited significantly greater structural diversity (i.e., distinct arrangements (sequence or order) and counts of repeat motifs) than EUR in Cohort 1, and than AMR, EAS, and EUR in Cohort 2 (Fig. 4a,b). Cohort 2 also showed more unique repeat structures, which may reflect both its larger sample size and more noise from lower sequencing depth, with the *RFC1* locus displaying the highest structural heterogeneity (Fig. 4c,d).

**Fig. 4.**
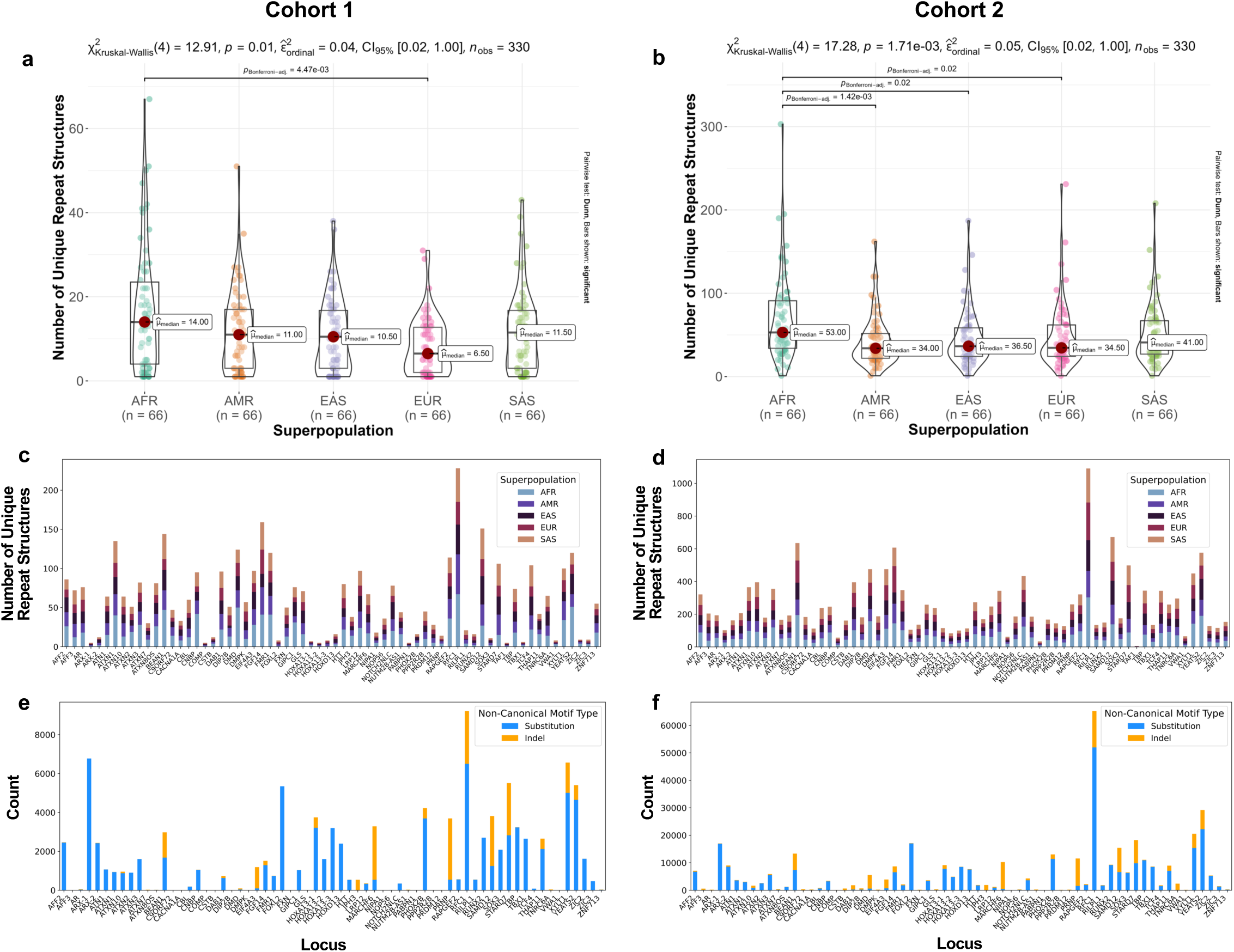
Unique repeat structures and non-canonical motifs in disease-associated TR loci (a– f). Annotated violin–boxplots with raw data overlay showing the number of unique repeat structures identified across 66 disease-associated TR loci in each superpopulation (AFR, AMR, EAS, EUR, SAS) for Cohorts 1 (**a**) and 2 (**b**). Statistically significant differences in the number of unique repeat structures between superpopulations, assessed by the Kruskal–Wallis test, are indicated above the plots. Where the Kruskal–Wallis test was significant, pairwise comparisons of groups were conducted using Dunn’s test with Bonferroni correction, and significant pairwise differences are also indicated above the plots. Stacked bar plots illustrating the number of unique repeat structures across all disease-associated TR loci for each superpopulation in Cohorts 1 (**c**) and 2 (**d**). Stacked bar plots displaying counts of substitution- and indel-based non-canonical motifs in disease-associated TR loci for Cohorts 1 (**e**) and 2 (**f**).

Substitution-based non-canonical motifs preserving reading frames were more frequent in coding TRs, whereas indel-based non-canonical motifs were more frequent in non-coding TRs (Fig. 4e,f). The most frequent non-canonical motifs were consistent across cohorts and populations, often recurring at the same position within the repeat tract and shared across ≥3 populations, providing increased confidence in the validity of these flagged non-canonical motifs (Supplementary Fig. 2).

Collectively, these findings reveal extensive locus- and population-specific TR diversity, characterized by alleles and length-dependent interruption patterns uniquely enriched in specific populations, underscoring their role in modulating repeat stability and epidemiological variation in RE disorders.

### Clinical relevance of disease-associated TR alleles

Mean frequencies of normal and full-mutation alleles were approximately 95% and ≤0.37%, respectively, across all loci in these two cohorts. No significant differences in mean frequencies were observed between superpopulations across loci (Supplementary Fig. 3a–f).

In addition to the 225 full-mutation alleles that were found across 17 loci, we identified 55 reduced-penetrance alleles across five loci, 13 premutation alleles across five loci, and 779 intermediate alleles across four loci. Although several of these allelic classes were distributed across ≥3 populations (Fig. 5a), many REs exhibited ancestry-specific distributions, including REs of *ATXN10* in AMR, *ZIC3* in EUR (notably Finnish), and *RUNX2* and *NOTCH2NLC* in EAS, consistent with prior observations^24–27^. In contrast, *FGF14* REs were detected in all populations except AFR.

**Fig. 5.**
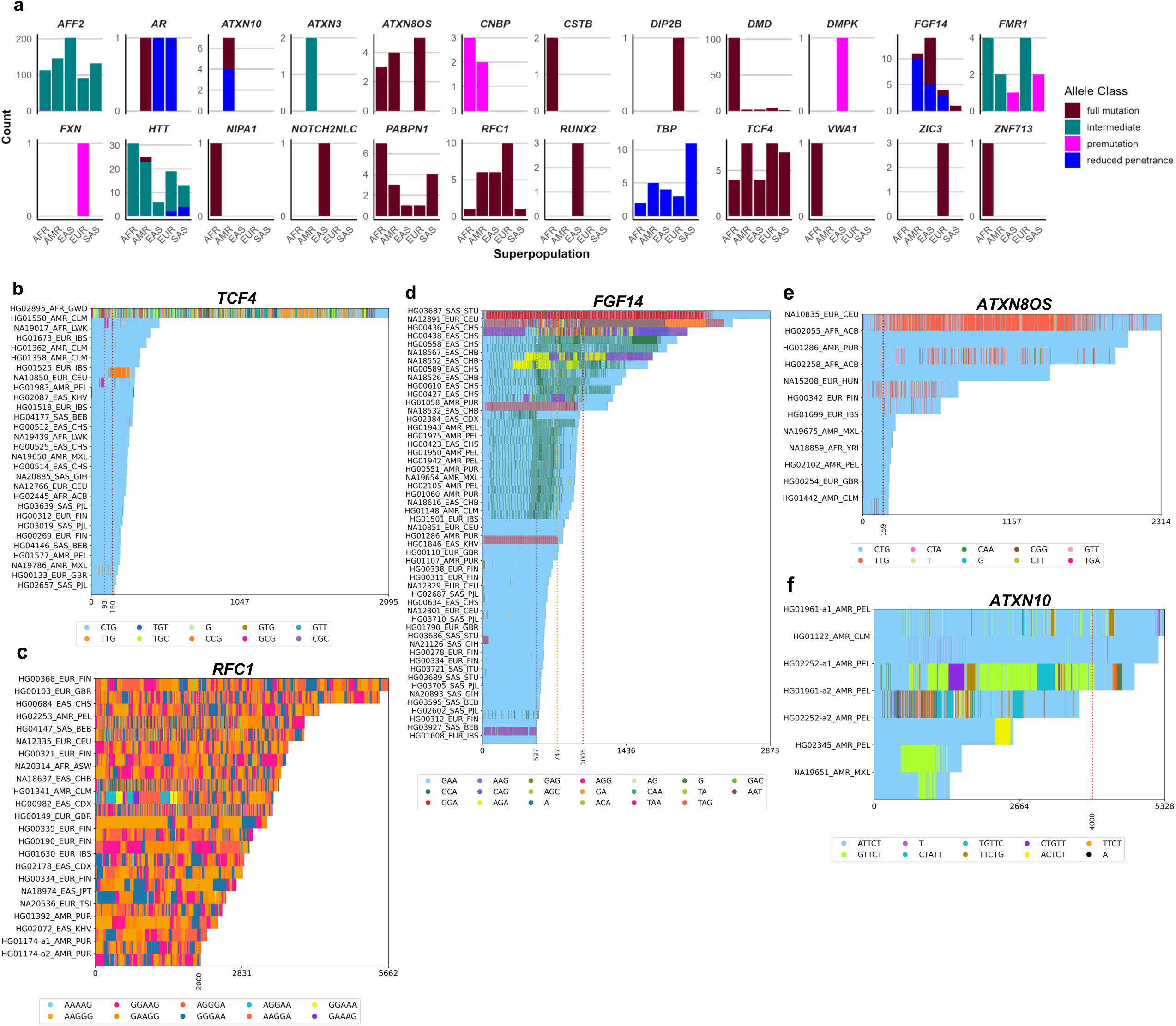
Disease-associated TR alleles, their counts, and sequence compositions (a–f). Counts of intermediate, premutation, reduced-penetrance, and full-mutation alleles across superpopulations (AFR, AMR, EAS, EUR, SAS) in Cohorts 1 and 2 (**a**). Sequence compositions of expanded *TCF4* (**b**), *RFC1* (**c**), *FGF14* (**d**), *ATXN8OS* (**e**), and *ATXN10* (**f**) alleles. Individual IDs, superpopulation, and population codes are shown for each TR sequence (*y*-axis: sample identifiers, and superpopulation and population information; *x*-axis: repeat length in bp). Sequences are sorted by length, longest on top. The canonical motif is shown in light sky blue; each non-canonical motif is assigned a distinct color. Legends (bottom of each panel) list the ten most frequent non-canonical motifs detected by TRMotifAnnotator. Vertical dotted lines indicate reported repeat size thresholds: gray for the upper bounds of normal, orange for the lower bounds of premutation alleles, and red for full-mutation alleles.

Among disease-associated TRs, *DMD* full mutations were most prevalent (111/225), consistent with prior reports^1^, occurring predominantly (92%) in AFR and less frequently in other populations (Fig. 5a). Most *DMD* full mutations exhibited non-canonical motifs, typically at the 3’ end (Supplementary Fig. 4a). Given that only uninterrupted alleles have been associated with Duchenne muscular dystrophy^28^, and *DMD* full mutations are common in the general population, the disease association of interrupted alleles remains uncertain.

*TCF4* full mutations, the second most frequent (31/225), occurred at similar rates in AFR/EAS and AMR/EUR/SAS. Approximately 50% of *TCF4* full mutations were comprised of pure CTGs, while interrupted motifs were more frequent in AFR (48%) and EUR (≤41%) than AMR, EAS, and SAS (0.34–12%) (Fig. 5b). Several novel non-canonical motifs (e.g., CCG, GCG) were evident in *TCF4* expansions, and differences in interruption types/patterns may underlie ancestry- and individual-specific variation in Fuchs endothelial corneal dystrophy risk, warranting further study.

Although numerous additional REs were detected, only a subset likely confer disease risk or higher penetrance based on sequence composition. Large alleles at pentanucleotide TR loci mostly involved expansions of canonical or benign motifs. At *RFC1*, 22 heterozygous carriers of the pathogenic AAGGG motif were observed across all populations, and one AMR individual was homozygous for the biallelic expansion associated with CANVAS; these sequences exhibited high structural complexity (Fig. 5c). At *FGF14*, most reduced-penetrance and full-mutation alleles exhibited 19–91% non-canonical motif content, with uniquely shared repeat structures enriched for GGA/GCA motifs in AMR and EAS (Fig. 5d), consistent with the non-pathogenicity of interrupted *FGF14* REs in SCA27A^21^. At *ATXN8OS*, 12 full mutations were identified; five had pure CTG repeats, while others showed up to 17% non-canonical content in AMR and up to 58% in EUR, including one EUR individual with four CGG interruptions associated with increased SCA8 risk and earlier onset^29^ (Fig. 5e). *ATXN10* REs in AMR (including two compound heterozygotes) harbored novel non-canonical motifs (e.g., GTTCT, ACTCT) of unknown significance with up to 67% non-canonical content (Fig. 5f).

Collectively, we identified 28 heterozygous full mutation alleles associated with pediatric or adult-onset recessive RE disorders (*AR*, *CSTB*, *RFC1*, *VWA1*, *ZIC3*), including one individual homozygous for an *RFC1* expansion consistent with CANVAS. Additionally, 67 individuals carried dominant full mutation alleles associated with adult-onset disorders (*HTT*, *NIPA1*, *NOTCH2NLC*, *PABPN1*, *TCF4*, *FGF14*, *ATXN8OS*, *ATXN10*), while six individuals carried dominant full mutation alleles associated with pediatric-onset disorders (*DIP2B*, *RUNX2*, *ZIC3*, *ZNF713*). Together, individuals harboring alleles conferring potential disease risk comprised 6% of the cohorts—when *DMD* and interrupted (non-pathogenic) *FGF14* REs were excluded.

LRS further revealed methylation, mosaicism, and X-inactivation patterns relevant for pathogenicity assessment. At *DIP2B*, a single full mutation in an EUR individual exhibited marked mosaicism (200–1200 CCG repeats) without the hypermethylation associated with gene silencing (Supplementary Fig. 4b). At *ZIC3*, full mutation allele in a female exhibited skewed X-inactivation of the expanded allele (Supplementary Fig. 4c). These findings highlight the importance of LRS-enabled comprehensive TR assessment to accurately evaluate true disease-associated full-mutation alleles.

### Local ancestry patterns of disease-associated TRs

At most disease-associated TR loci, the inferred local ancestry (based on a 500 kb window centered on the TR) matched the reported superpopulation in ∼70% of AFR, EAS, and EUR individuals. In contrast, only 35–39% of AMR and SAS individuals carried matching or EUR ancestry (Supplementary Fig. 5a). Several loci deviated markedly from this pattern, revealing pronounced locus-specific ancestry enrichment. *NOTCH2NLC* exhibited nearly universal EUR ancestry (90%) even among AFR individuals. Conversely, *PHOX2B* and *RAPGEF2* carried EAS ancestry in ∼30% of EUR individuals, while *ZIC3* carried EUR ancestry in ∼30–50% of EAS and SAS individuals.

To better understand the genomic context of these loci, we analyzed ±2.5 Mb flanking regions, which revealed contiguous or fragmented ancestry tracts around TRs. At *NOTCH2NLC*, a broad EUR-enriched segment spanned the TR in all superpopulations (Supplementary Fig. 5b), indicating an extended EUR background. In contrast, *PHOX2B*, *RAPGEF2*, and *ZIC3* showed discrete ancestry switches at the TR (Supplementary Fig. 6a–c), consistent with recombination events or locus-specific ancestry shifts due to admixture. Notably, *PHOX2B* and *RAPGEF2* also displayed EAS-enriched flanks in AMR, EUR, and SAS individuals.

Inferred local ancestries of premutation, reduced-penetrance, and full-mutation alleles revealed additional patterns (Fig. 6,a–e). In AFR and EUR individuals, local ancestry generally matched the superpopulation. In AMR individuals, however, *ATXN10* reduced-penetrance and full-mutation alleles aligned with AMR ancestry, consistent with the known ancestral origin of *ATXN10* REs^30,31^. By contrast, reduced-penetrance *TBP* alleles were distributed across multiple ancestries (AFR, EAS, EUR), and *FGF14* reduced-penetrance and full-mutation alleles were enriched for AMR or EUR ancestry in AMR, EUR, and SAS individuals but spread across ancestries (except AFR) in EAS, indicating locus- and population-specific ancestry contributions.

**Fig. 6.**
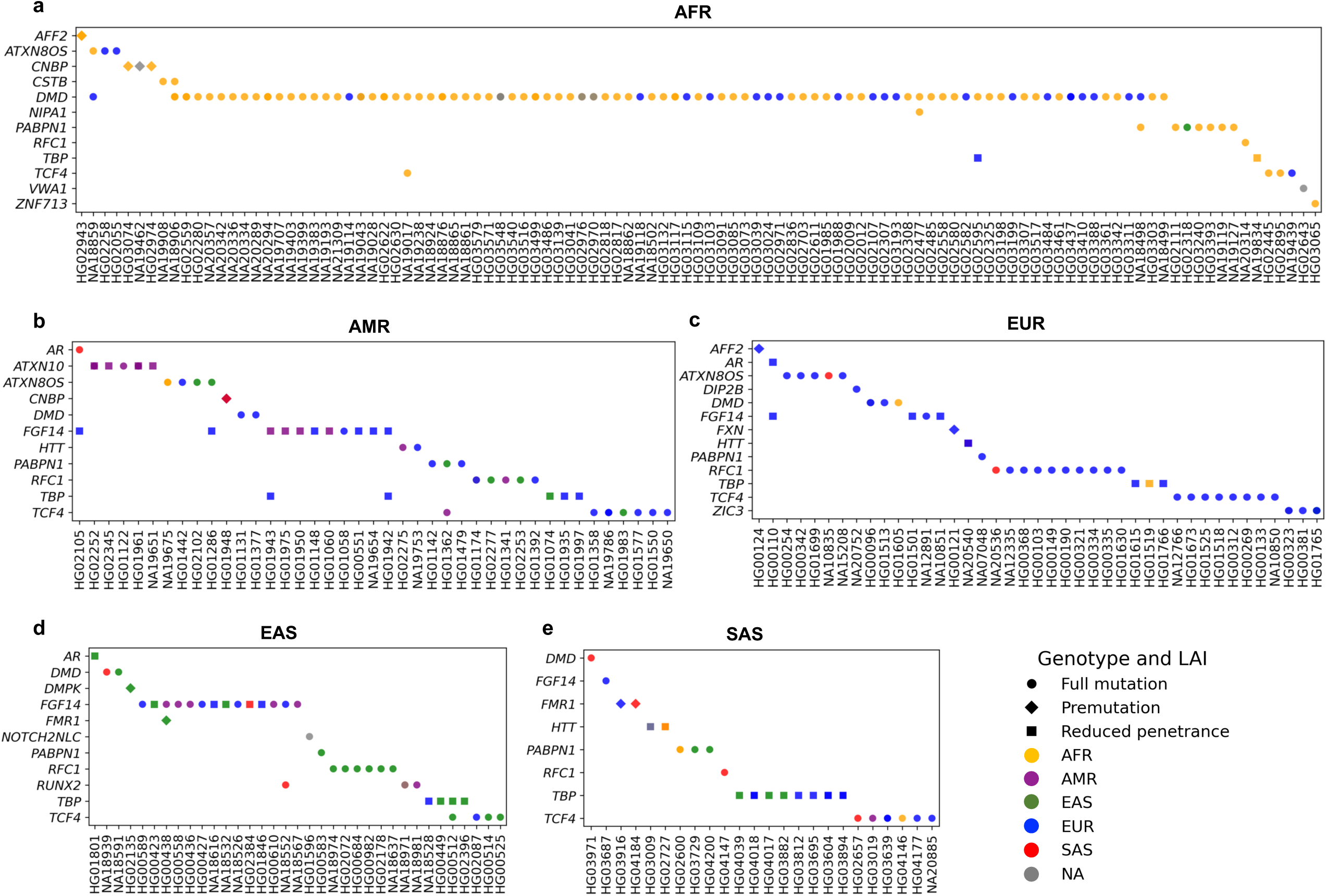
Local ancestry inference of premutation, reduced-penetrance, and full-mutation TR alleles (a–e). Local ancestry inference of disease-associated TR loci with premutation, reduced penetrance, and full mutation alleles, grouped by the superpopulation (AFR (**a**), AMR (**b**), EUR (**c**), EAS (**d**), SAS (**e**)). Each panel shows individuals (*x*-axis) and disease-associated TR loci (*y*-axis). Data point shape indicates allele class (premutation, reduced penetrance, or full mutation), and color denotes inferred local ancestry. Samples lacking intersecting RFMix calls at the TR are labeled “NA”. For homozygotes, the ancestry of one haplotype is shown.

TR sequence composition was also associated with ancestry. Nearly pure reduced-penetrance *FGF14* alleles (<1% non-canonical content) in EUR and EAS individuals were observed exclusively on EUR ancestry backgrounds, while highly interrupted full-mutation alleles (>49% non-canonical content) mapped to EUR or AMR ancestry in EAS, SAS, and EUR individuals. Full mutations with intermediate non-canonical content (19–47%) in EAS and AMR individuals were more broadly distributed, highlighting the influence of ancestral background on sequence purity and interruption patterns. Similarly, the TGG-interrupted *TCF4* 16–17 repeat alleles, most frequent in AMR and observed in other populations, consistently mapped to EUR/AMR local ancestry across AMR, EUR, and SAS individuals (Supplementary Fig. 7a), consistent with a haplotype likely shared among these populations, rather than being specific to AMR.

Together, these results illustrate that disease-associated TRs often reside on population-specific haplotypes shaped by admixture, exhibiting locus- and motif-specific ancestry patterns that diverge from individuals’ overall superpopulation. These localized ancestry switches may reflect the diverse historical and demographic dynamics driving TR variation and disease risk, though some may result from local ancestry misassignment.

### Phylogenetic relationships of disease-associated TRs

Phylogenetic analysis of TR sequences in the high-coverage Cohort 1 data revealed locus- and population-specific divergence, reflecting a complex interplay of conservation and diversification. Principal component analysis of tree summary statistics (summarized in Additional Data Table 1) separated deeply branched, unbalanced loci (e.g., *RFC1*, *FGF14*), indicative of rapid diversification, from compact, conserved loci in coding, untranslated, or promoter regions (e.g., *ATXN7*, *DMPK*), consistent with stronger purifying selection (Fig. 7a).

**Fig. 7.**
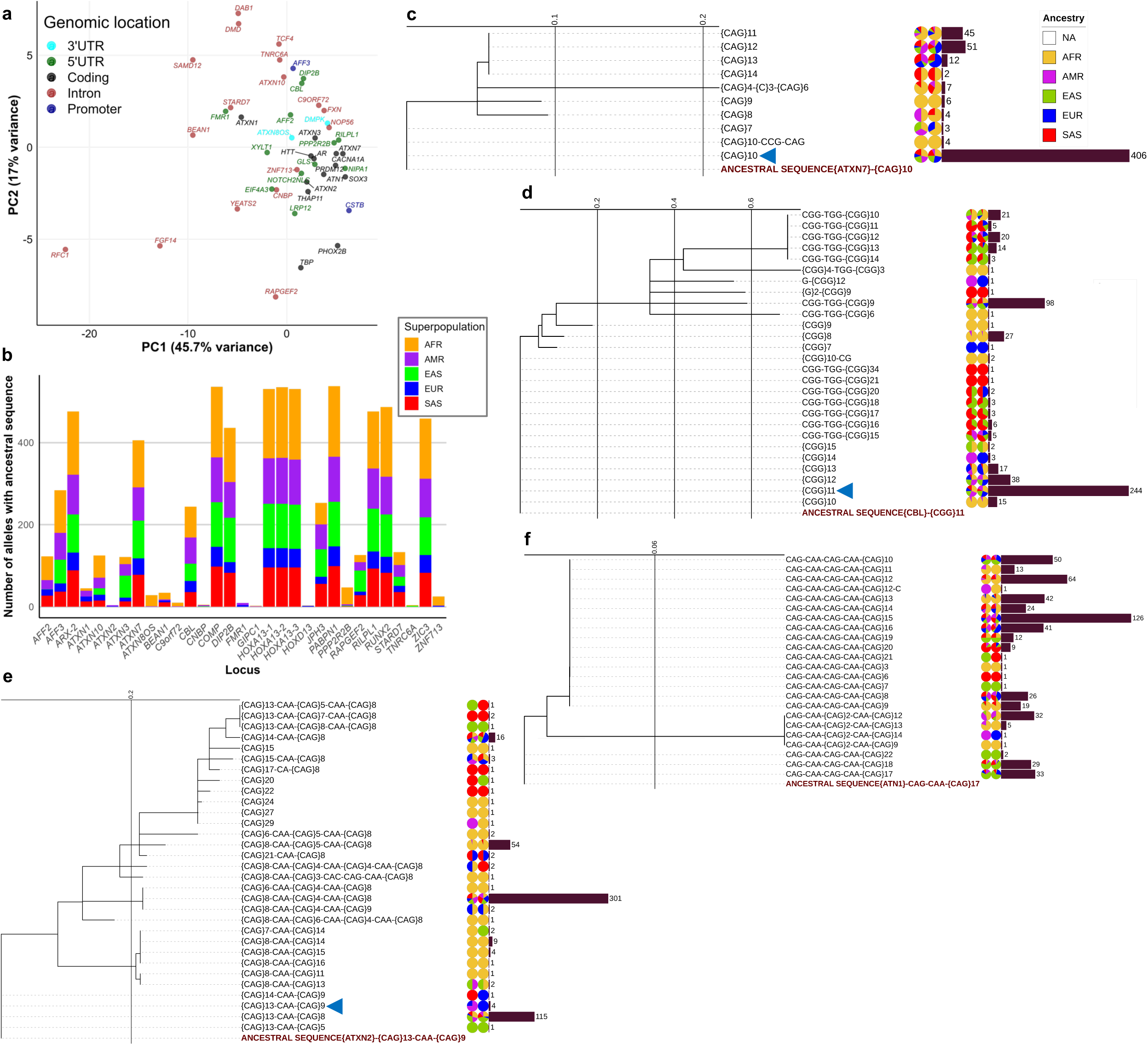
PCA of phylogenetic tree statistics and representative trees of disease-associated TR loci (a–f). Principal component analysis of summary statistics from phylogenetic trees (**a**). Stacked bar plot showing counts of alleles at 31 loci retaining the ancestral allele (**b**). Phylogenetic trees of *ATXN7* (**c**), *CBL* (**d**), *ATXN2* (**e**), and *ATN1* (**f**), based on unique repeat structures and rooted with the ancestral sequence (bottom). Branch lengths are shown on the internal scale of each panel. Repeat structures are labeled at the tips; haplotypes retaining the ancestral sequence are marked with a blue triangle. For each repeat structure, the first pie chart shows the proportion of alleles by superpopulation; the second pie chart shows the proportion of RFMix-inferred local ancestry. Adjacent bars indicate allele counts by repeat structures. Colors for superpopulations and inferred local ancestries are defined in the legend (**c**) and consistently applied to both.

At 31 loci, the ancestral sequence (derived from multiple alignments of seven primate genomes) was retained in at least one haplotype, and at 19 of these it was present in ≥100 (of 544) haplotypes across all superpopulations (Fig. 7b). This widespread retention likely reflects functional constraints and selective pressures that regulate TR stability and mutation dynamics. High retention was observed at *COMP*, *HOXA13*, *PABPN1*, and *ATXN7*. At *ATXN7*, the ancestral (CAG)_10_ appeared in 406 haplotypes, with other common derived variants carrying 1–3 additional repeats (Fig. 7c). At *CBL*, the ancestral (CGG)_11_ persisted in 244 haplotypes, alongside a common derived CGG-TGG-(CGG)_9_ variant (98 haplotypes), with longer CGG-TGG-(CGG)_11-34_ alleles enriched in EAS and SAS and shorter (CGG)_8_ alleles enriched in AFR (Fig. 7d). In contrast, at 12 loci the ancestral sequence was rare (<50 haplotypes) or absent, consistent with elevated mutation rates and/or a selective advantage of derived configurations. At *ATXN2*, for example, the ancestral (CAG)_13_-CAA-(CAG)_9_ appeared in only four haplotypes, whereas the most frequent allele (301 haplotypes) had two CAA interruptions, (CAG)_8_-CAA-(CAG)_4_-CAA-(CAG)_8_, with a slightly modified variant, (CAG)_8_-CAA-(CAG)_5_-CAA-(CAG)_8_, enriched in AFR (Fig. 7e). Similarly, at *ATN1*, the ancestral CAG-CAA-(CAG)_17_ was completely absent, with all sequences carrying two CAA interruptions in a CAG-CAA-CAG-CAA-(CAG)_8-21_ configuration (Fig. 7f), potentially conferring greater stability and reducing disease risk. See Additional Data Fig. 3 for phylogenetic trees of all loci.

At most loci, the most frequent TR allele was shared across superpopulations, with structurally similar sequences clustering together (Supplementary Table 4). However, several loci exhibited population-specific divergence in sequence composition, which may contribute to ancestry-related differences in TR stability. At *ATXN1*, variants with a single CAT interruption formed a distinct clade from the ancestral and other sequences with two CAT interruptions, with one such variant, (CAG)_11_-CAT-(CAG)_16_, enriched in EAS (Supplementary Fig. 7b). Similarly, at *ATXN3*, sequences with CAA-AAG interruptions formed a separate clade from those with an additional CAA interruption (CAA-AAG-CAG-CAA), with (CAG)_2_-CAA-AAG-(CAG)_15_ predominant in EAS and SAS (Supplementary Fig. 7c).

These population-specific variants in *ATXN1* and *ATXN3* exhibited increased sequence purity (a feature associated with higher instability and disease risk), while other loci showed population-specific non-canonical motifs or elevated mutation rates. This includes the large impure tracts in *ATXN10* enriched in AMR (Supplementary Fig. 7d) and *BEAN1* alleles with 52– 57 repeats harboring distinctive non-canonical motifs in AFR (Supplementary Fig. 8). In contrast, *GIPC1* sequences with GGC-GGA-GGG-AGC interruptions formed a distinct clade at the tips of the tree, yet consistently observed across all superpopulations (Supplementary Fig. 9), indicative of a recent configuration subject to ongoing diversification.

Collectively, these findings reveal the influence of functional constraints, demographic history, and locus-specific mutation dynamics on sequence purity and structural divergence at disease-associated TR loci.

### Population-wide haplotype patterns surrounding disease-associated TRs

Recombination-driven haplotype structures are critical in determining the imputation accuracy of REs, TR stability, and disease risk. We profiled regional haplotype patterns around disease-associated TRs across populations, benchmarking them against local recombination signatures.

Consistent with genome-wide patterns^8,39^, principal component analysis of phased single-nucleotide variants (SNVs) within 1 Mb of each TR showed that the top three components explained ∼80% of the variance, clearly separating superpopulations (Fig. 8a) and highlighting the impact of population history on haplotypic backgrounds and the need for population-specific TR analyses.

**Fig. 8.**
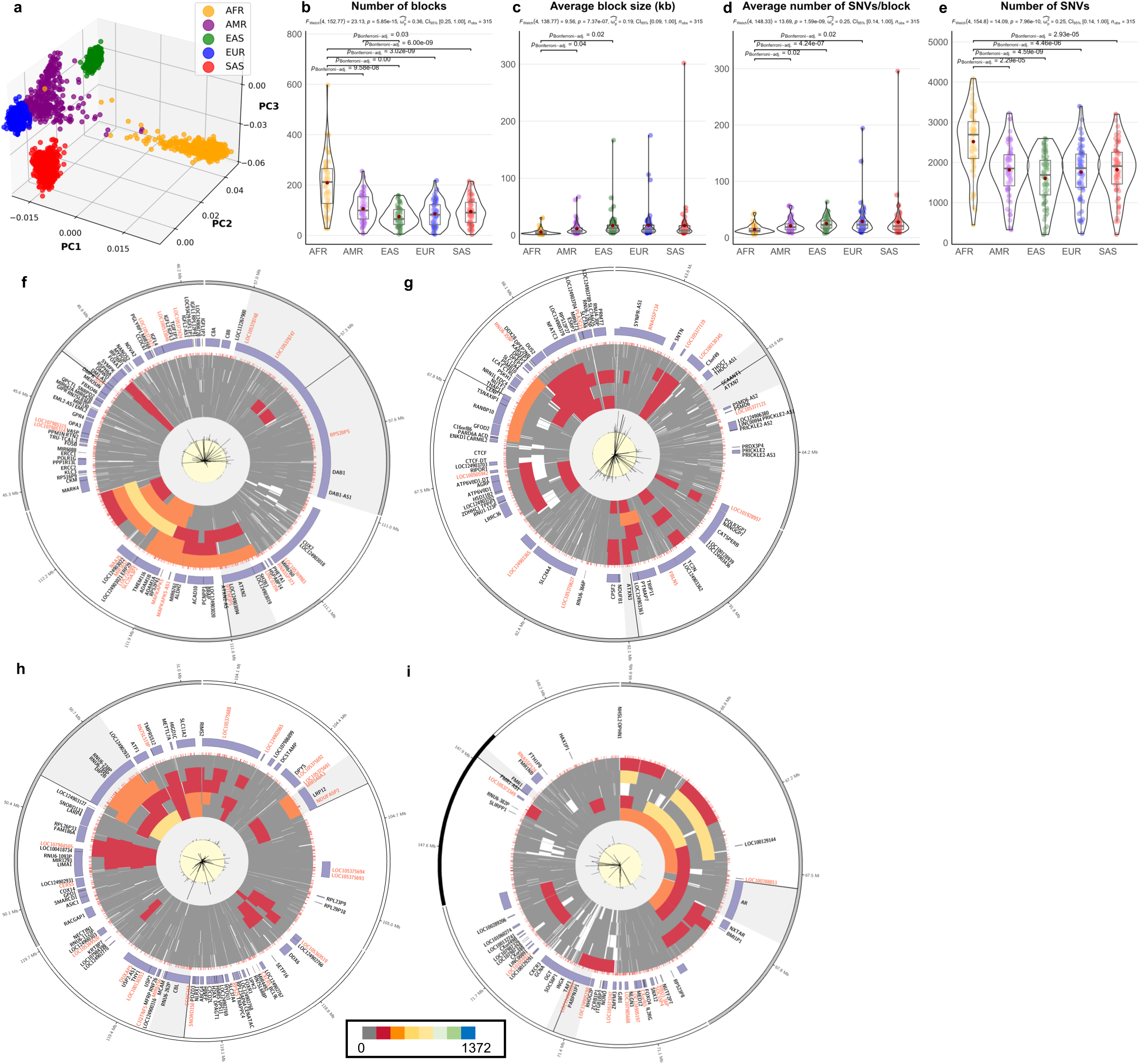
Linkage disequilibrium (LD) blocks around disease-associated TR loci (a–i). Principal component analysis of bi-allelic single-nucleotide variants from a 1 Mb region encompassing 66 disease-associated TRs, based on short-read sequencing data from 2,504 unrelated individuals in the 1kGP (**a**). Colors indicate the reported superpopulations of the individuals. LD statistics (**b–e**). Statistically significant differences in LD blocks between superpopulations were assessed using one-way ANOVA, with post-hoc unpaired pairwise *t*-tests (Bonferroni-adjusted, *p*) to identify significant pairwise differences. Significant pairwise differences in the number of LD blocks (**b**), the average LD block size (in kilobases) (**c**), the average number of SNVs per LD block (**d**), and the total number of SNVs between superpopulations (**e**) are shown; the red central point indicates mean. Circos plots (**f–i**) display, from outermost to innermost: the ideogram (Layer 1), gene track (Layer 2), and repeat track of simple repeats (Layer 3). Layers 4–8 show LD blocks for each superpopulation: AFR, AMR, EAS, EUR, and SAS. Average recombination rates from 1kGP and deCODE Genetics (scale range: 1–100 cM/Mb) are shown in Layers 9 (light gray background) and 10 (light yellow background), respectively. All tracks are visualized across a 1-Mb region encompassing selected genes with disease-associated TRs. Gray highlights indicate the gene of interest, while black highlights within the gray mark the TR location. 1-Mb regions around loci shown are: *DAB1*, *ATXN2*, and *DMPK* (**f**); *ATXN7*, *ATXN3*, and *THAP11* (**g**); *DIP2B*, *LRP12*, and *CBL* (**h**); and *AR*, *TAF1*, and *FMR1* (**i**). The color legend at the bottom corresponds to the number of SNVs (range: 0–1372) in the LD block heatmaps (Layers 4–8).

LD analysis showed that AFR individuals had more, smaller LD blocks, each with fewer SNVs, consistent with higher recombination and diversity (Fig. 8b–e). Across populations, we observed a significant inverse correlation between average LD block size and the number of recombination hotspots defined as regions with recombination rates exceeding 1 cM/Mb (Supplementary Fig. 10a), reinforcing the critical role of recombination in shaping the haplotype architecture around disease-associated TRs. Of the 66 loci studied, 27 lay within 10 kb of hotspots on both flanks, while 35 were more isolated (Supplementary Fig. 10b,c).

See Additional Fig. 4 for LD and recombination signatures across TR loci, stratified by chromosome. Haplotype structures varied widely: large, dense LD blocks at loci such as *ATXN2*, *THAP11*, and *AR* coincided with recombination deserts (regions lacking a hotspot within 100 kb of the TR) (Fig. 8f–i). Conversely, loci including *CACNA1A* and *GIPC1* exhibited fragmented LD patterns indicative of recombination-active regions, while *ATXN10* and *MARCHF6* exhibited localized recombination suppression within broader recombination-active contexts (Supplementary Fig. 11a,b).

LD landscapes surrounding disease-associated TRs are provided in Supplementary Figs. 12 and 13 and summarized in Supplementary Tables 5 and 6. In populations except AFR, several TRs (e.g., *DMPK*, *TBP*) were spanned by contiguous LD blocks, reflecting relatively conserved haplotypes outside of AFR. Conversely, more TRs (e.g., *CACNA1A*, *SOX3*) resided near LD block boundaries or at junctions between adjacent blocks across populations. Notably, in some cases, no LD blocks span the TR in specific populations (e.g., *PRNP* in EAS, *SOX3* across nearly all populations, and *FOXL2* in both EAS and EUR). These patterns reflect regional differences in LD structure and imply that imputation accuracy for TR alleles may vary substantially across populations and loci.

Local ancestry switches observed at *PHOX2B*, *RAPGEF2*, and *ZIC3* (Supplementary Fig. 6a–c) correspond closely with LD block patterns; TRs at these loci were typically positioned near edges of LD blocks, consistent with recombination activity and ancestry transitions influencing haplotype structure.

Evidence of shared haplotypes between populations was apparent at several loci (Supplementary Table 6). For example, *ATXN10* exhibited nearly identical regional LD block structures in AMR and EUR (Supplementary Fig. 14). Further, SNVs (minor allele frequency ≤ 0.05) co-segregating with REs highlighted tagging potential: the SNV rs41524547-G, previously associated with reduced-penetrance and full mutation alleles^31^, occurred on all AMR carriers of expanded *ATXN10* alleles in our assemblies. Among 16,328 gnomAD individuals with TR genotypes, 16 (14 AMR and 2 AFR) carried *ATXN10* alleles >50 repeats; rs41524547-G was present in all but one AMR carrier. Local ancestry analysis of these 16 individuals confirmed predominantly AMR ancestry, including the two AFR individuals who were assigned AMR local ancestry at the locus. Notably, both bi-allelic RE carriers were homozygous for this SNV (Fig. 9), reinforcing rs41524547-G’s utility as a tagging SNV for imputing *ATXN10* REs, particularly within populations maintaining strong LD between the SNV and the TR.

**Fig. 9.**
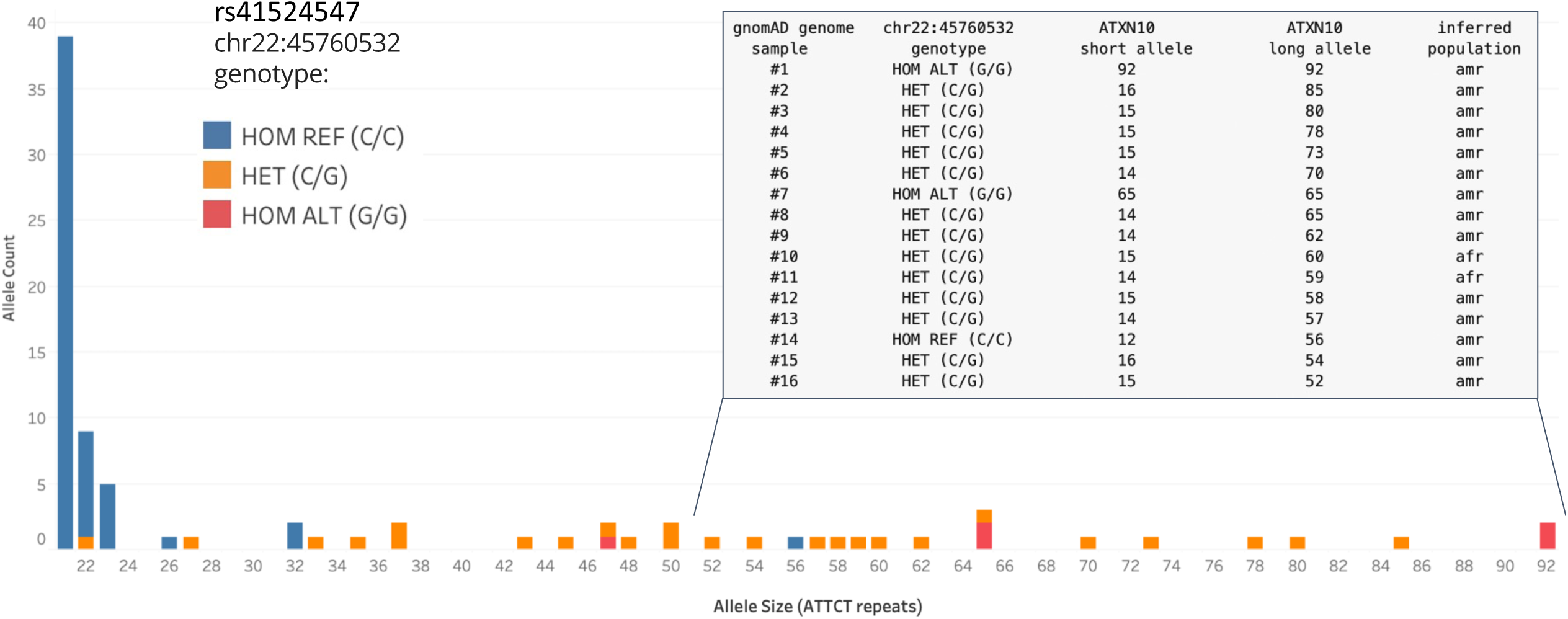
***ATXN10* repeat size distribution and association with rs41524547 genotype in gnomAD genomes.** Histogram of *ATXN10* allele size distribution (≥21 repeats) in 16,328 gnomAD genomes, colored by rs41524547 (GRCh38: chr22:45760532) genotype. The inset table reports the repeat and SNV genotypes for samples with exceptionally long alleles (>50 repeats), illustrating that expansions >32 repeats almost always co-occur with the non-reference G allele at rs41524547.

These findings demonstrate heterogeneity in haplotype structures at disease-associated TR loci, reflecting locus-specific genomic features and population-specific evolutionary history, with important implications for RE imputation, interpretation of disease associations, and the design of haplotype-based analyses in diverse populations.

## Discussion

We present the first comprehensive LRS-based analysis of disease-associated TRs, revealing allele-, locus-, and population-specific variations underlying epidemiological disparities in RE disorders. Such variation is exemplified at the *DIP2B* locus, where stable alleles were less prevalent in EUR, consistent with the population specificity of the associated disorder^19^ (Fig. 2a). At *FGF14*, EAS showed a narrow distribution (5–50 repeats) but a higher frequency of very large (>180) and intermediate (180–250) alleles, which could predispose to disease. However, compared to other populations, most large EAS alleles had significantly higher non-canonical base content due to interruptions, which mitigate pathogenicity and align with the lower prevalence of SCA27B in this population^21^ (Fig. 2b; Fig. 10). In contrast, EUR and SAS individuals carried more large, uninterrupted alleles (50–200), which are prone to instability and expansion, consistent with higher disorder prevalence in these groups^32^. LRS-based profiling classified 87% of long *FGF14* alleles as interrupted and likely benign, while delineating population-specific distributions of unstable alleles that mirror RE disorder prevalence. These findings underscore the importance of understanding repeat copy distributions for identifying at-risk populations and demonstrate the utility of LRS-based TR analysis in carrier screening and population risk assessment.

**Fig. 10.**
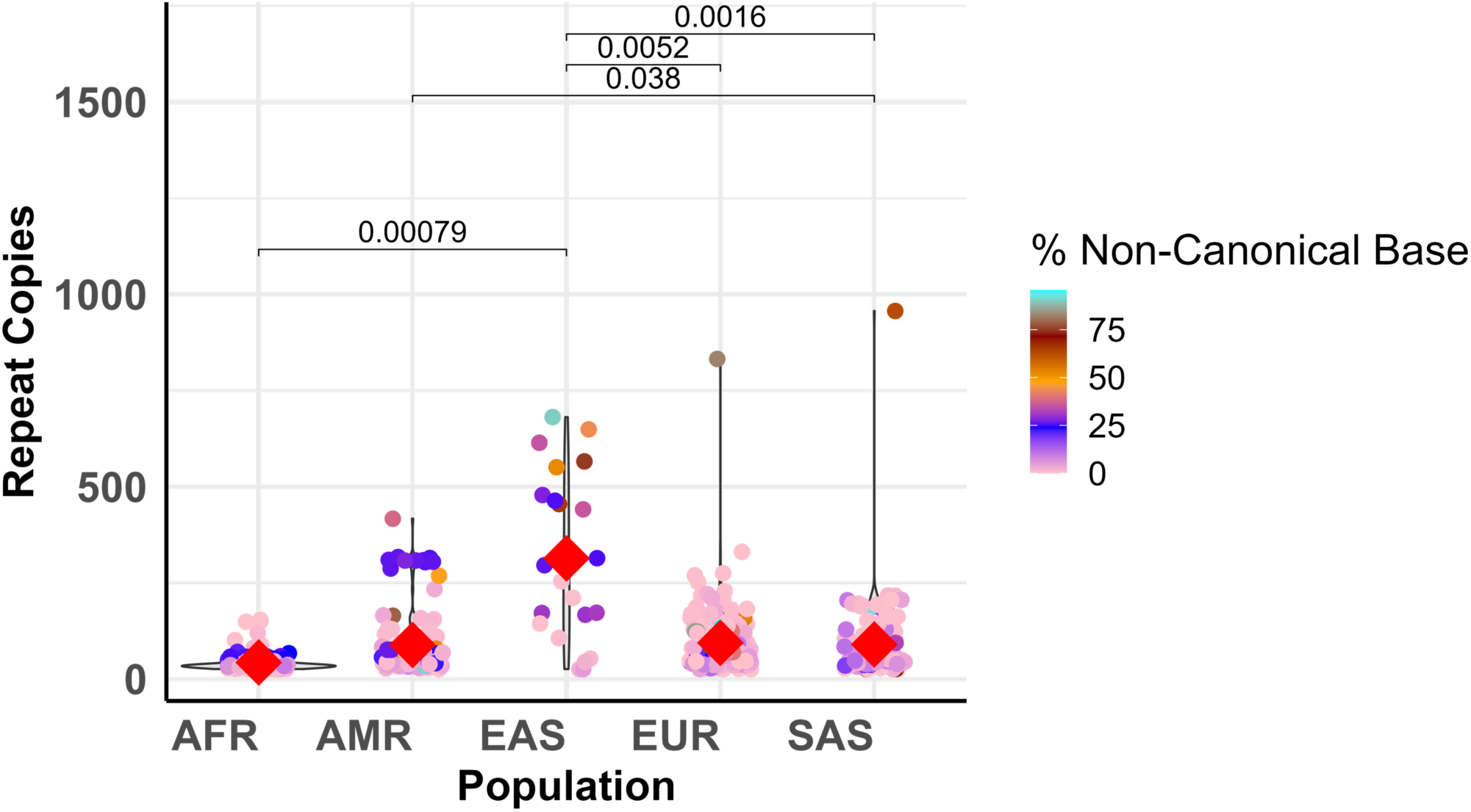
Distribution of *FGF14* repeat copies and non-canonical base content by population. Violin plot shows the distribution of repeat copy numbers (>25 copies) across five superpopulations. Each point represents an individual allele, colored by its percentage of non-canonical base content (pink to cyan gradient). Average non-canonical base content for each population is indicated by a red triangle within the violin. Pairwise comparisons of non-canonical base content between populations were performed using Wilcoxon rank-sum tests with Bonferroni correction; significant differences are annotated above with adjusted *p*-values.

TR size- and ancestry-stratified allele frequency data from large, diverse populations are essential for interpreting RE–disease associations^3^. Without such context, alleles rare in one population may be misclassified as pathogenic if they are common in underrepresented groups. For example, the *DMD* RE, previously implicated in Duchenne muscular dystrophy, is rare in most populations but too frequent in AFR to support pathogenicity. Analysis of the larger Cohort 2 revealed greater allelic diversity than Cohort 1, underscoring the need for large, ancestrally diverse datasets to fully capture RE variation and improve pathogenicity inference.

Sequence-informed analysis revealed 6% of individuals in the general population carry fully expanded alleles associated with potential disease risk. These expansions often exhibit ancestry-specific patterns and are largely linked to adult-onset disorders. While these individuals were presumed asymptomatic, some REs may manifest later in life. Notably, six individuals carried dominant alleles for pediatric-onset RE disorders without symptoms, raising questions about the reported pathogenicity and penetrance of these expanded alleles. This includes the X-linked *ZIC3* expansion^24^—previously implicated in a reportedly male-lethal disorder—identified in three apparently unaffected individuals (two females and one male). These findings suggest more REs presumed to be fully penetrant may, in fact, show incomplete penetrance or variable expressivity.

While demonstrating the power of LRS to resolve TR variation, these findings also highlight the ethical challenges of incidental findings in population-scale studies, where disclosure may be impractical, undesired, or beyond the scope of consent. Population-wide screening must balance these concerns with the need for genetic counseling and the implications of predictive testing in asymptomatic individuals, particularly when identifying carriers of severe RE disorders like Huntington disease^33^.

Our observation of distinctive motif patterns within TR alleles of defined length ranges, such as at *BEAN1*, supports that specific sequence compositions are associated with particular genotypes, with TR diversity differing between populations, highest in AFR individuals, and often featuring non-canonical motifs or interruptions shared across individuals^34^. More broadly, the locus- and population-specific evolutionary trajectories uncovered in this study show how TR characteristics, functional constraints, and demographic history contribute to allelic diversity and the emergence of unstable alleles. Differences in sequence purity, motif structure, and conservation across populations may reflect distinct selective pressures and mutational dynamics that drive variation in disease risk and prevalence.

Previous local ancestry investigation at disease-associated TR loci relied on broad genomic windows (5 Mb) and short-read sequencing, limiting resolution to smaller REs^3^. In contrast, our analysis of LRS data—which enables more accurate haplotype phasing—TR genotyping, and sequence composition analysis, focused on a narrower 500-kb region and revealed locus- and allele-specific local ancestry patterns for TRs, including larger REs such as those at the *FGF14* and *ATXN10* loci. These findings refine and extend prior work by providing finer-scale insights into local haplotype structure across diverse populations.

We further show that disease-associated TRs are embedded within complex, locus- and population-specific genetic backgrounds influenced by selection, recombination, admixture, and local ancestry—often differing from genome-wide ancestry. These patterns highlight the need to interpret REs within their full genomic and ancestral context. The frequent placement of TRs at LD block boundaries or recombination-active regions suggests their susceptibility to haplotype fragmentation, with important implications for imputation, genetic screening, and the transferability of association signals across populations. These insights emphasize that accurate risk prediction and carrier identification require population-aware approaches that account for local ancestry patterns and haplotype diversity, especially in understudied and genetically diverse populations.

By integrating sequence-resolved TR data from LRS with ancestry-aware genomic analysis, our work provides a foundational resource for interpreting TR variation in its full evolutionary, demographic, and genomic context. This integrated approach enables more accurate delineation of both global and population-specific disease risks, facilitates the identification of ancestry-restricted alleles and motif configurations, informs the refinement of pathogenicity thresholds, and supports improved clinical interpretation of REs. As LRS data of diverse populations are generated, integrating detailed sequence, motif, genetic, and local ancestry data will be essential for accurately defining risk alleles, haplotypes, and their prevalence. Our findings also underscore the need to move beyond single-population reference databases and simple allele size cutoffs toward a multidimensional, sequence-, and context- aware interpretation of RE disorders.

## Data Availability

All data produced in the present work are contained in the manuscript. Additional data files are available at https://doi.org/10.6084/m9.figshare.30337696.

https://humanpangenome.org/data/

https://millerlaboratory.com/1KGP-LRS.html#Data

https://www.internationalgenome.org/data-portal/data-collection/hgsvc2

https://ftp.1000genomes.ebi.ac.uk/vol1/ftp/data_collections/1KG_ONT_VIENNA/

https://doi.org/10.6084/m9.figshare.30337696

## Methods

### Study cohorts and haplotype-resolved genome assemblies

We analysed 2,526 haplotypes from 1,263 healthy individuals, assembled from PacBio or Nanopore LRS data across two cohorts. The first cohort (*n*=272) comprised ∼35–40× genomes from three publicly available datasets^12,16,17^, as described by Chiu *et al*^13^. The second cohort (*n*=1019) consisted of ∼15× Nanopore data from 1kGP individuals sequenced by Noyvert *et al*^18^. For Cohort 2, we generated haplotype-resolved assemblies for each individual. After excluding four samples due to read extraction failure and 24 overlapping with the first cohort, 991 genomes from the second cohort were retained for analysis.

### TR genotype and sequence composition analysis

Disease-associated TR loci were identified in the assemblies following Chiu *et al*^13^, recording the assembly scaffold, coordinates, and strand in BED format for each locus. We used Tabix to query each locus, selecting the closest overlapping TR by minimal start-end difference. TR sequences were extracted with Pysam and genotyped using Tandem Repeats Finder^35^ with parameters used in Straglr^36^. In parallel, disease-associated TR loci were genotyped in alignments using Straglr in targeted mode. Assembly-derived TR genotypes that showed zygosity discrepancies relative to Straglr alignment-based genotypes, likely due to insufficient read coverage, were excluded from analysis. The remaining genotypes were classified by repeat copy number and/or sequence composition using thresholds and repeat motifs curated from Rajan-Babu IS *et al*^4^ and other sources^1,37–39^.

Disease-associated TR sequences were further analysed with TRMotifAnnotator, a Python tool that identifies both canonical motifs and non-canonical variant motifs resulting from substitutions or indels within the canonical motif. TRMotifAnnotator parses assembled sequences of TR tracts, splitting them at canonical motif occurrences to identify intervening non-canonical motifs. Its output includes (1) a TSV file with repeat genotypes, sequence structures, and motif counts, and (2) a color-coded plot where each track represents a sequence and colors denote different repeat motifs.

### Local ancestry, phylogenetic, and linkage disequilibrium analyses

Local ancestries at disease-associated TR loci were inferred from SNVs called on each of the 2,526 assembled haplotypes aligned to GRCh38. Phased VCFs containing bi-allelic SNVs from 500-kb regions flanking each TR were extracted; ∼90% of these windows contained ≥300 SNVs, providing sufficient marker density. We applied RFMix^40^ to these VCFs following the approach described by Ibañez *et al*^3^.

Rooted phylogenetic trees for each disease-associated TR locus were constructed by aligning TR sequences representing unique repeat structures with predicted ancestral TR sequences. The ancestral TR sequences were derived from multiple alignments of seven primate genomes^41^ (<u>Ensembl</u>) using Clustal Omega^42–44^. Trees were visualized with iTOL^45^, and summary statistics were computed with *treestats*^46^.

For principal component and LD analyses, we used phased SNV calls from 2504 unrelated individuals in <u>1kGP phase 3</u>. Principal components were computed from SNVs within 1-Mb window flanking each TR locus using PLINK. For LD analysis, phased SNVs from 5-Mb windows were processed to generate population-specific DET files, which were then used to analyse LD patterns at disease-associated TR loci.

## Data and code availability

All supporting data are included within the article and its Supplementary Information. Additional data files generated in this study are available at https://doi.org/10.6084/m9.figshare.30337696.

Haplotype-level TR genotypes, sequence composition data, summary statistics, and inferred local ancestries are available via REDatlas, an interactive web application that also visualizes the geographical distribution of RE disorders, based on curated data from GeneReviews, OMIM, and recent literature. REDatlas is accessible at https://atlasred.streamlit.app, and its source code is available at https://github.com/wf-TRs/REDatlas.

Detailed command-line workflows and parameters used for genome assembly, variant calling, local ancestry inference, and phylogenetic and LD analyses are provided in the Supplementary Methods. The TRMotifAnnotator tool is available at https://github.com/wf-TRs/TRMotifAnnotator.

## Acknowledgments

This study was supported by the Canadian Institutes of Health Research (CIHR) Project Grant to J.M.F, I.B, and I.S.R.B. (PJT-169074). I.S.R.B. is a recipient of the CIHR Research Excellence, Diversity, and Independence (REDI) Early Career Transition Award (DI2-190730). We thank the Digital Research Alliance of Canada for resource allocation (ynp-672-aa) and The University of British Columbia Advanced Research Computing for allocations that supported data generation and storage. We are thankful to the Human Pangenome Reference Consortium, the 1000 Genomes Project ONT Sequencing Consortium, Human Genome Structural Variation Consortium, and Noyvert *et al* for their efforts and access to the datasets analysed in the study.

## Author contributions

I.S.R.B. conceived, designed, and executed the study, performing the data and figure generation, analysis, and interpretation. R.C. contributed to genome assembly generation, repeat sequence extraction, and manuscript review. B.W. contributed to data generation from gnomAD samples. I.C. contributed to REDatlas creation with guidance and input from I.S.R.B. I.B. and J.M.F. contributed to experimental design guidance, data interpretation, and manuscript review. I.S.R.B. led manuscript writing and overall project coordination.All authors have read and approved the manuscript.

## Statistical information

Statistical comparisons of TR copy distributions across superpopulations (AFR, AMR, EUR, EAS, SAS) and study cohorts were conducted using the Kolmogorov–Smirnov test to assess differences in repeat copy number distributions at each of the 66 known disease-associated TR loci. To control for multiple testing, Bonferroni correction was applied, and loci with adjusted *p*-values below the significance threshold (0.05) were considered significant. Comparisons of unique repeat structure counts between study cohorts were evaluated using the Kruskal–Wallis test, followed by Dunn’s pairwise tests with Bonferroni adjustment. Allelic frequency comparisons across superpopulations employed one-way ANOVA with Games–Howell post-hoc tests, also adjusted for multiple comparisons. LD block statistics around TR loci were analyzed by one-way ANOVA, with post-hoc unpaired pairwise t-tests and Bonferroni correction used to identify differences in LD block number, size, and SNV density across superpopulations. PCA summarized patterns in phylogenetic tree statistics and bi-allelic SNVs within 1 Mb regions encompassing TR loci. Pairwise comparisons of non-canonical base content between superpopulations at the *FGF14* locus were performed using Wilcoxon rank-sum tests with Bonferroni correction for multiple testing. Significant differences were reported based on adjusted *p*-values.

## Supplementary information

Supplementary information is included in this manuscript.

Correspondence and requests for materials should be addressed to [Indhu-Shree Rajan-Babu].

### Supplementary Figure Legends

**Supplementary Fig. 1 Allelic classifications and repeat length distributions of known disease-associated TR loci in Cohort 1.**

Raincloud plots show repeat length distributions across 66 known disease-associated TR loci, derived from *de novo* haplotype-resolved genome assemblies. The *y*-axis represents the estimated number of repeat copies. Genotypes are stratified by assigned ancestry or superpopulation (AFR, AMR, EUR, EAS, SAS) of the 272 individuals from Cohort 1. Scatter plot colors indicate the allelic class of each genotyped TR allele: normal, intermediate, premutation, unknown, reduced penetrance, or full-mutation.

**Supplementary Fig. 2 Non-canonical motifs in disease-associated TR loci shared between superpopulations and allelic classes (a,b).**

Heatmaps show the most frequent non-canonical motifs within and between the different allelic classes (Normal, NL; Intermediate, IM; Premutation, PM; Unknown, UNK; Reduced Penetrance, RP; Full-mutation, FM) at each disease-associated TR locus, shared across individuals of different superpopulations. Results from Cohort 1 (**a**) and Cohort 2 (**b**) are shown. Repeat loci are shown on the *y*-axis, and allele classes on the *x*-axis. Heatmap color indicates the number of superpopulations that share the same most frequent non-canonical motif. Cells without annotation or “.” indicate that the most frequent pattern is a pure repeat of the canonical motif. Longer non-canonical motifs (>10 base pairs) are represented as “…”. Uncolored cells marked with “.” indicate allelic classes that are either not present (e.g., no UNK class at *FMR1*) or were not observed in the respective cohort (e.g., no premutations at *AFF2* in Cohort 1).

**Supplementary Fig. 3 Allelic frequencies of disease-associated TRs (a–f).**

Annotated violin–boxplots with raw data overlay showing the frequencies (%) of normal (**a**), intermediate (**b**), premutation (**c**), unknown (**d**), reduced penetrance (**e**), and full-mutation (**f**) alleles in Cohorts 1 and 2 across all loci. Comparisons of mean allelic frequencies across populations are shown. Group differences were tested using one-way ANOVA, and pairwise comparisons were performed using the Games–Howell test with Bonferroni correction. The *n* values indicate the number of disease-associated TR loci studied.

**Supplementary Fig. 4 Sequence composition and methylation profiles of expanded alleles at disease-associated TR loci (a–c).**

Sequence composition of expanded alleles at the *DMD* locus (**a**). Individual IDs, superpopulation, and population codes are indicated for each TR sequence (*y*-axis), sorted by repeat length (*x*-axis, in base pairs (bp)) with longer alleles at the top. The canonical motif is highlighted in light sky blue; each identified non-canonical motif is shown in a unique color. The bottom legend lists the top ten non-canonical motifs detected by TRMotifAnnotator. Vertical dotted gray and red lines indicate the upper bound of normal allele length and the lower bound of full-mutation allele length, respectively. Methylation profiles of normal and expanded *DIP2B* alleles in NA/GM20752 (**b**). IGV screenshot of the *DIP2B* promoter region showing methylation status around the expanded allele (top) and the normal haplotype (bottom). Multiple insertion signatures suggest repeat-length mosaicism, with observed allele sizes of ∼600, 750, 900, 1500, and 3600 bp. Bases in red indicate methylated CpGs; blue bases indicate unmethylated CpGs. The (CGG)_n_ TR and promoter positions are annotated below the panel. Methylation profiles of normal and expanded *ZIC3* alleles in HG00323 (**c**). IGV screenshot showing the expanded haplotype with 12 repeats (top) and the normal haplotype (bottom). Bases in red indicate methylated CpGs; blue bases indicate unmethylated CpGs. The TR position is annotated below the panel.

**Supplementary Fig. 5 Local ancestries of disease-associated TRs (a,b).**

Local ancestry proportions within the 500 kb window encompassing each disease-associated TR locus, inferred by RFMix and stratified by superpopulation (**a**). Stacked bar plots show the proportions of AFR, AMR, EAS, EUR, SAS, or Unknown local ancestries (*y*-axis) at each TR locus (*x*-axis) in Cohort 1 (left) and Cohort 2 (right), with colors corresponding to local ancestry codes shown in the legend. Local ancestry in the 5 Mb region flanking the *NOTCH2NLC* TR, stratified by superpopulation (**b**). The top subpanel of each superpopulation shows stacked bar plots of local ancestry proportions (*y-*axis) across consecutive 500 kb windows within the 5 Mb region (*x-*axis). The bottom subpanel of each superpopulation shows a tile plot of local ancestry for each haplotype from Cohorts 1 and 2. The red dotted vertical line in the bottom panel marks the TR start position.

**Supplementary Fig. 6 Local ancestries in the 5 Mb region spanning *PHOX2B*, *RAPGEF2*, and *ZIC3* TRs (a–c).**

Local ancestry across the 5 Mb region surrounding the *PHOX2B* (**a**), *RAPGEF2* (**b**), and *ZIC3* (**c**) TRs, stratified by superpopulation. The top panel in each subfigure shows stacked bar plots of local ancestry proportions (*y-*axis) across consecutive 500 kb windows within the 5 Mb region (*x-*axis). The bottom panel shows a tile plot of local ancestry for each haplotype from Cohorts 1 and 2. The red dotted vertical line in the bottom panel marks the TR start position.

**Supplementary Fig. 7 Inferred local ancestry of 16–17-repeat *TCF4* alleles and phylogenetic trees of *ATXN1*, *ATXN3*, and *ATXN10* repeat sequences (a–d).**

Stacked bar plot showing local ancestry proportions of 16**–**17-repeat *TCF4* alleles with TGG interruptions, compared to all other alleles across superpopulations (**a**). Phylogenetic trees of *ATXN1* (**b**), *ATXN3* (**c**), and *ATXN10* (**d**), based on unique repeat structures and rooted with the ancestral sequence (bottom). Branch lengths are shown on the internal scale of each panel. Repeat structures are labeled at the tips. For each repeat structure, the first pie chart shows the proportion of alleles by superpopulation; the second pie chart shows the proportion of RFMix-inferred local ancestry. Adjacent bars indicate allele frequency by repeat structures. Colors for superpopulations and inferred local ancestries are defined in the legend and consistently applied to both.

**Supplementary Fig. 8 Phylogenetic tree of *BEAN1* repeat sequence.**

**Supplementary Fig. 9 Phylogenetic tree of *GIPC1* repeat sequence.**

**Supplementary** Fig. 10 **Recombination hotspots around disease-associated TR loci (a–c).**

Scatter plots depict the relationship between mean linkage disequilibrium block size (kb) and the number of recombination signals (>1 cM/Mb; 1kGP) within 1 Mb of each locus across five superpopulations (AFR, EUR, SAS, EAS, AMR) (**a**). Points represent loci, the red line shows the linear regression fit, and the Pearson correlation coefficient (r) and *p*-value are indicated in each panel. A similar trend was noted in deCODE (data not shown). Heatmaps showing distances from each TR midpoint to the nearest recombination hotspots (>1 cM/Mb) in 1kGP (**b**) and deCODE (**c**) datasets. Loci are listed on the *y*-axis; distances to the nearest 5’ and 3’ hotspots are shown for each TR. Heatmap cells display distances (in base pairs), with colors indicating recombination signal intensity as per the legend.

**Supplementary Fig. 11 Linkage disequilibrium blocks around disease-associated TR loci (a,b).**

Circos plots display, from outermost to innermost: the ideogram (Layer 1), gene track (Layer 2), and repeat track of simple repeats (Layer 3). Layers 4–8 show linkage disequilibrium blocks for each superpopulation: AFR, AMR, EAS, EUR, and SAS. Average recombination rates from 1kGP and deCODE Genetics (scale range: 1–100 cM/Mb) are shown in Layers 9 (light gray background) and 10 (light yellow background), respectively. All tracks are visualized across a 1-Mb region encompassing selected genes with disease-associated TRs. Gray highlights indicate the gene of interest, while black highlights within the gray mark the TR location. 1-Mb regions around loci shown are: *CACNA1A*, and *GIPC1* (**a**), and *ATXN10*, and *MARCHF6* (**b**). The color legend at the bottom corresponds to the number of SNVs (range: 0–1372) in the LD block heatmaps (Layers 4–8).

**Supplementary Fig. 12 Linkage disequilibrium block patterns around disease-associated TR loci.**

Each panel displays linkage disequilibrium block patterns and SNV density in the genomic regions flanking disease-associated TR loci across superpopulations (AFR, AMR, EAS, EUR, SAS). Horizontal colored segments show defined linkage disequilibrium blocks for each population, with color intensity indicating SNV density as shown in the accompanying color bar. *X*-axis shows ∼100-kb window around each TR locus. The vertical dashed red line marks the exact position of the TR locus.

**Supplementary Fig. 13 Linkage disequilibrium block patterns around disease-associated TR loci (contd..).**

Panels show a continuation of Supplementary Fig. 12 with the remaining TR loci.

**Supplementary Fig. 14 Linkage disequilibrium block patterns and SNV maps at the *ATXN10* TR locus.**

Circos plots depicting the genomic context of the *ATXN10* TR region on chromosome 22. The outermost layer (Layer 1) shows the chromosome ideogram, followed by the gene track (Layer 2), SNV track showing rsIDs or genomic coordinates of SNVs in linkage disequilibrium block (Layer 3), and repeat annotation track for simple repeats (Layer 4). Layers 5–14 show the SNVs within linkage disequilibrium block and linkage disequilibrium block structure from PLINK for each superpopulation (AFR, AMR, EAS, EUR, SAS). Average recombination rates from 1kGP and deCODE Genetics (scale range: 1–100 cM/Mb) are shown in the innermost layers in light gray and yellow backgrounds, respectively. The color scale at the bottom indicates the number of SNVs (ranging from 0 to 1372) in linkage disequilibrium block heatmaps.

### Supplementary Table Legends

**Supplementary Table 1 Population differences in TR copy number distributions in Cohort 1.**

Kolmogorov-Smirnov tests were conducted to compare repeat copy number distributions across superpopulations for each disease-associated TR locus in Cohort 1. Bonferroni correction was applied to account for multiple comparisons, with loci exhibiting significant differences identified accordingly. Both unadjusted and adjusted *p*-values are presented.

**Supplementary Table 2 Population differences in TR copy number distributions in Cohort 2.**

Kolmogorov-Smirnov tests were conducted to compare repeat copy number distributions across superpopulations for each disease-associated TR locus in Cohort 2. Bonferroni correction was applied to account for multiple comparisons, with loci exhibiting significant differences identified accordingly. Both unadjusted and adjusted *p*-values are presented.

**Supplementary Table 3 Cohort comparison of TR distributions.**

Kolmogorov-Smirnov tests compared repeat copy number distributions between Cohort 1 and Cohort 2 for each disease-associated TR locus and superpopulation. Bonferroni correction was applied to account for multiple comparisons, and loci with significant differences were identified. Both unadjusted and adjusted *p*-values are reported.

**Supplementary Table 4 Repeat structures of disease-associated TRs in Cohort 1, and their frequencies by reported** superpopulation and by RFMix-inferred local ancestry.

**Supplementary Table 5 Linkage disequilibrium block patterns surrounding disease TR loci.**

Linkage disequilibrium blocks overlapping disease TR loci, their start and end coordinates of the linkage disequilibrium block along with the number of SNVs per linkage disequilibrium block in each superpopulation. Linkage disequilibrium blocks with more than 100 SNVs are highlighted.

**Supplementary Table 6 SNVs in linkage disequilibrium shared between superpopulations.**

For each disease-associated TR locus, the number and percentage of SNVs in linkage disequilibrium shared between the compared superpopulations are shown along with the rsIDs of the shared SNVs in the last column.

