## Supplementary Methods for "Population-scale disease-associated tandem repeat analysis reveals locus and ancestry-specific insights"

**Affiliations:**
[1]Department of Medical Genetics, The University of British Columbia, Vancouver, BC, Canada
[2]Canada's Michael Smith Genome Sciences Centre, BC Cancer, Vancouver, BC, Canada
[3]Program in Medical and Population Genetics, Broad Center for Mendelian Genomics, Broad Institute of MIT and Harvard, Cambridge, MA, USA
[4]Center for Genomic Medicine, Massachusetts General Hospital, Harvard Medical School, Boston, MA, USA
[5]BC Children's Hospital Research Institute, Vancouver, BC, Canada

**Corresponding author:**
Indhu-Shree Rajan-Babu, Ph.D.
Department of Medical Genetics
The University of British Columbia
4500 Oak Street, Vancouver, BC V6H 3N1, Canada

### Supplementary Methods

#### Genome assembly and haplotype phasing (Cohort 2)

For the second cohort (*n*=1,019), Nanopore reads were assembled using `Flye (v2.9.3)`, aligned back to the assembly with `minimap2 (v2.24)`, and sorted with `samtools`. Haplotype-resolved assemblies were generated with `HapDup (v0.12)`:

```
flye --nano-hq <assembly.fasta> --out-dir <sample_dir> –threads 32

minimap2 -ax map-ont -t 32 <assembly.fasta> <ont.fastq> | samtools view -bhS
- | samtools sort -@ 32 -m 4G - -o <aln.bam>

singularity exec --bind <sample_dir> + hapdup_0.12.sif hapdup --assembly
<assembly.fasta> --bam <aln.bam> --out-dir <hapdup_dir> -t 32 --rtype ont
```

#### TR analysis

Disease-associated TR loci were genotyped in alignments (BAM/CRAM) files using Straglr[1] in targeted mode:

```
      Straglr \
            <aln.bam or aln.cram> \
            <reference.fasta> \
            <straglr_output> \
            --loci <diseaseTR.bed> \
            --genotype_in_size \
            --min_support 4 \
            --min_cluster_size 2 \
            --max_num_clusters 2 \
            --nprocs 16 \
            --tmpdir <work_dir> \
            --include_partials
```

TR genotypes were stratified by superpopulation based on self-reported ancestry, geographic origin, and 1000 Genomes Project ancestry annotations. Repeat copy distributions were plotted with Python packages: `matplotlib`, `seaborn`, and `ptitprince`. Pairwise distributional differences in repeat copy number between superpopulations (within and across cohorts) were assessed at each locus using the Kolmogorov–Smirnov test, with $p$-values corrected for multiple testing by the Bonferroni method, and significance was defined as an adjusted $p < 0.05$.

Sequence compositions of disease-associated TRs were analysed with TRMotifAnnotator executed with the following command:

```
python TRMotifAnnotator.py \
    --input <sequence.fa> \
    --output <prefix> \
    --canonical-motif <canonical motif> \
    --max-mers <canonical motif length> \
    --vlines "[(value1, 'color1'), (value2, 'color2')]" \
    --locus <locus-name>
```

Differences in the number of unique repeat structures and allelic frequencies between superpopulations were evaluated using the `ggbetweenstats` R package. Kruskal–Wallis tests (with post hoc Dunn's test and Bonferroni correction) were used for repeat structure comparisons across superpopulations. Differences in the mean allelic frequencies across superpopulations were assessed using one-way analysis of variance (ANOVA), followed by Games–Howell tests with Bonferroni correction.

#### Local ancestry inference

Variants were called on each of the 2,526 assembled haplotypes aligned to GRCh38 using `minimap2` and `paftools.js`:

```
minimap2 -cx asm5 --cs <genome.fa> <haplotype.fa> | sort -k6,6 -k8,8n >
<output.paf>

paftools.js call -f <genome.fa> <output.paf> > <haplotype.vcf>
```

The resulting VCFs for each haplotype were merged per sample using `bcftools merge`. Genotypes were encoded as "0|1" or "1|0" for heterozygous variants present on one haplotype, and as "1|1" when present on both. Local ancestry inference was performed using `RFMix` (`v2.03-r0`) on phased bi-allelic single nucleotide variants within 500-kb windows flanking each disease-associated TR locus:

```
rfmix -f <phased.chr{$chr}.vcf.gz> \
      -r <1kGP_high_coverage_Illumina.chr{$chr}.phased.vcf.gz> \
      -m <sample.txt> \
      -g <genetic_map_hg38_chr{$chr}.txt> \
      --chromosome={$chr} \
      -o <rfmix-output> \
      -n 5 -e 1 -c 0.9 -s 0.9 -G 15 \
      --n-threads=16
```

RFMix output for each individual was intersected with disease-associated TR coordinates using `bedtools intersect`. Superpopulation codes from 1kGP Phase 3 individuals[2] were appended to facilitate comparison between the assigned and inferred local ancestries at each TR locus.

#### Phylogenetic tree construction

Disease-associated TR sequences, along with predicted ancestral sequences derived from seven primate genomes, were aligned using `Clustal Omega`[3–5]:

```
clustalo -i <sequence.fasta> -o <aligned_sequence.fasta>

clustalo -i <aligned_sequence.fasta> --guidetree-
out=<phylo_tree_ancestral.nwk>
```

Phylogenetic trees were visualized using `iTOL`[6], annotated with superpopulation, inferred local ancestry, and the frequencies of each TR allele. Summary statistics for each tree were computed using `treestats`[7] and subsequently analyzed by principal component analysis to reduce dimensionality and identify clustering patterns or shared structural features across loci.

#### Principal Component and Linkage Disequilibrium Analyses

Phased SNVs from 2,504 unrelated individuals from the 1kGP Phase 3 panel were extracted within 1-Mb windows surrounding each TR locus and converted to PLINK binary format for principal component analysis using PLINK (v2.00–10252019-avx2):

```
plink2 --vcf <merged.phased.1Mb.SNV.vcf> --make-bed --out
<merged.phased.1Mb.SNV>

plink2 --bfile <merged.phased.1Mb.SNV> --pca --out
<merged.phased.1Mb.SNV.pca>
```

The first three principal components (PC1, PC2, and PC3) were obtained from the resulting eigenvectors and visualized as a 3D scatter plot using Matplotlib.

For linkage disequilibrium (LD) analysis, SNVs within 5-Mb windows around each TR locus were extracted, annotated with rsIDs using `bcftools annotate`, and stratified by superpopulation. The annotated VCFs were converted to PLINK BED format, and LD blocks were identified using PLINK:

```
plink2 --vcf <merged.phased.5Mb.${pop}.vcf> --make-bed --out
<merged.phased.5Mb.${pop}> --threads 24 --update-sex <${pop}_sex.txt>

plink2 --bfile <merged.phased.5Mb.${pop}> --blocks no-pheno-req --blocks-max-
kb 500 --out <merged.phased.5Mb.${pop}.LD.blocks>
```

Differences in LD block characteristics between superpopulations were evaluated using one-way ANOVA, followed by post hoc unpaired pairwise *t*-tests with Bonferroni adjustment for multiple comparisons, implemented in the `ggstatsplot` R package. The following LD metrics were compared across superpopulations: number of LD blocks, mean LD block size (in kilobases), mean number of SNVs per LD block, and total number of SNVs within the region. Statistically significant pairwise differences ($p < 0.05$) were identified for each metric.

LD patterns and recombination signatures from the 1kGP and deCODE[8] studies were visualized across 1-Mb regions flanking each TR locus using `Circos`[9]. Pearson correlation coefficients were calculated to assess the association between LD block size and recombination rates (>1cM/Mb) derived from the 1kGP and deCODE datasets.
