## Supplementary Figures 1-14 for "Population-scale disease-associated tandem repeat analysis reveals locus and ancestry-specific insights"

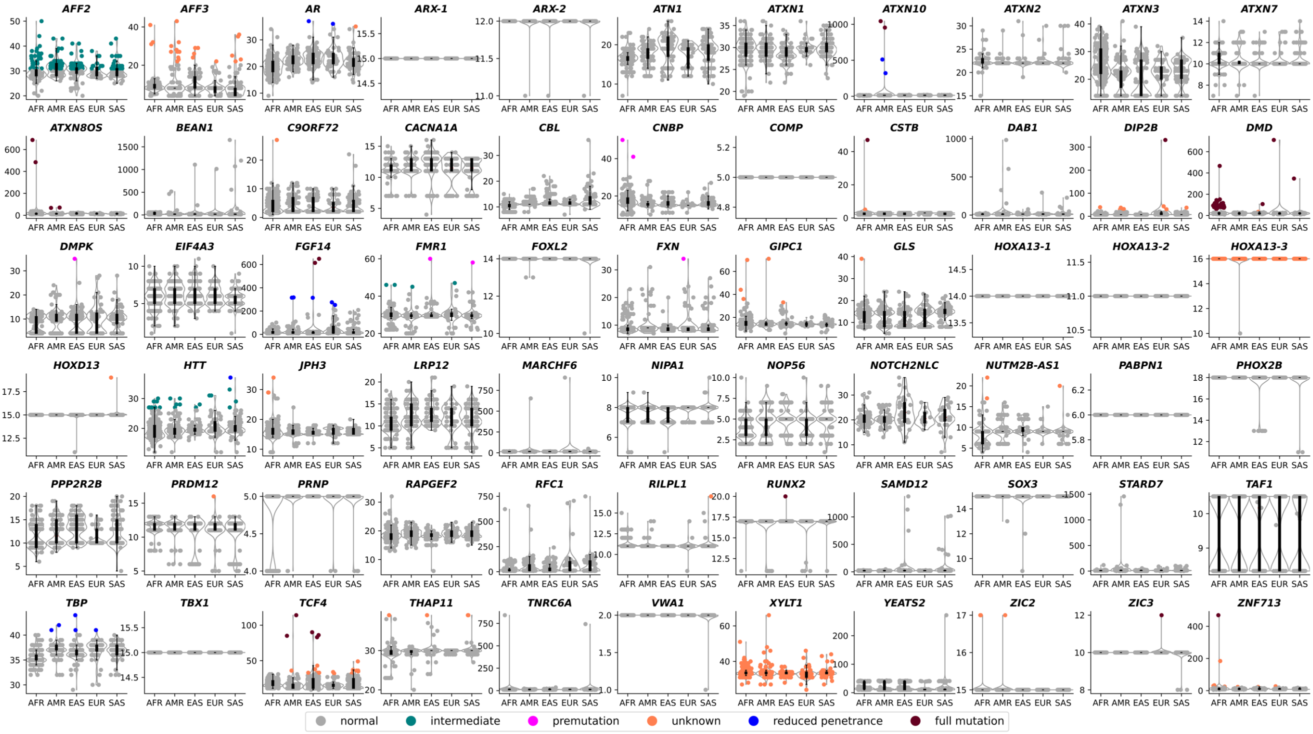


**Supplementary Fig. 1 | Allelic classifications and repeat length distributions of known disease-associated TR loci in Cohort 1.** Raincloud plots show repeat length distributions across 66 known disease-associated TR loci, derived from *de novo* haplotype-resolved genome assemblies. The *y*-axis represents the estimated number of repeat copies. Genotypes are stratified by assigned ancestry or superpopulation (AFR, AMR, EUR, EAS, SAS) of the 272 individuals from Cohort 1. Scatter plot colors indicate the allelic class of each genotyped TR allele: normal, intermediate, premutation, unknown, reduced penetrance, or full-mutation.

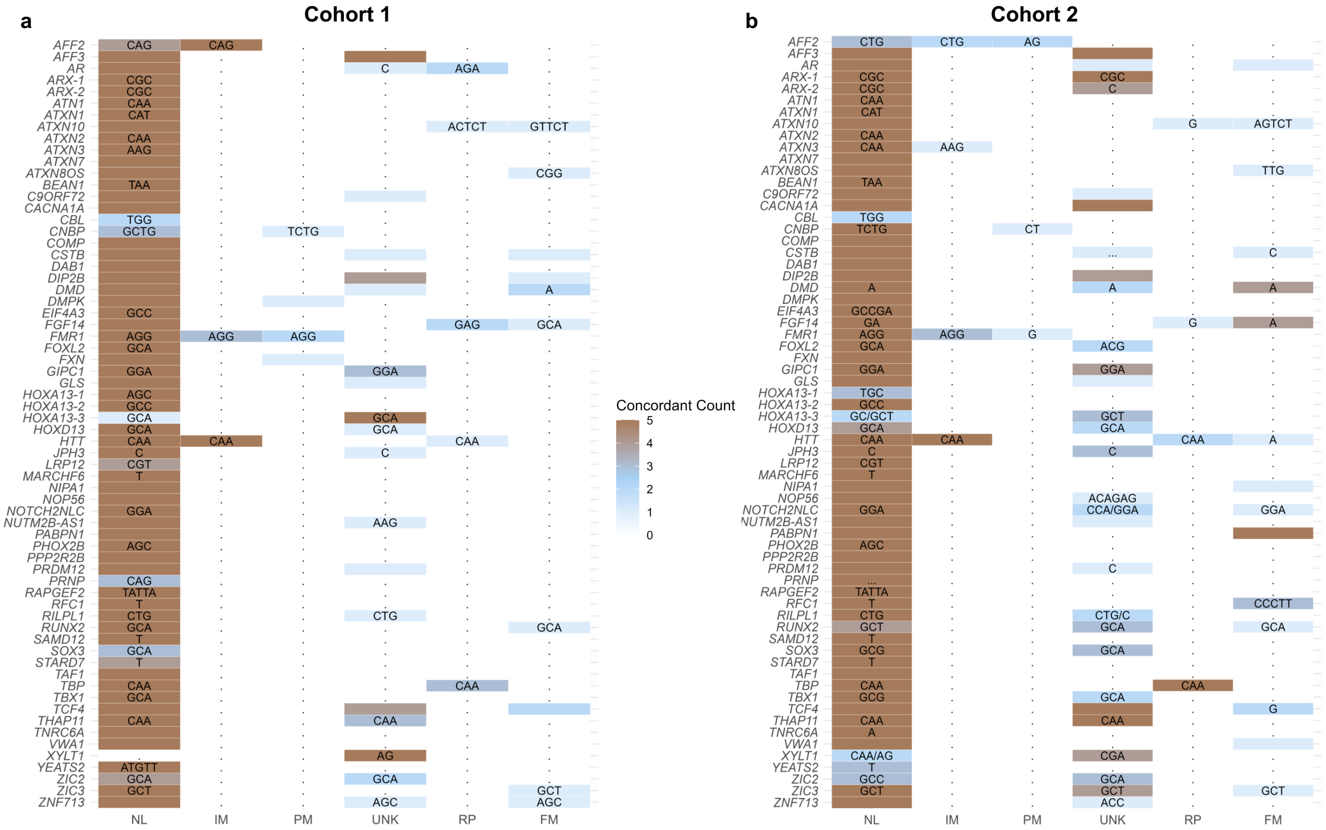


**Supplementary Fig. 2 | Non-canonical motifs in disease-associated TR loci shared between superpopulations and allelic classes (a,b).** Heatmaps show the most frequent non-canonical motifs within and between the different allelic classes (Normal, NL; Intermediate, IM; Premutation, PM; Unknown, UNK; Reduced Penetrance, RP; Full-mutation, FM) at each disease-associated TR locus, shared across individuals of different superpopulations. Results from Cohorts 1 (**a**) and 2 (**b**) are shown. Repeat loci are shown on the *y*-axis, and allele classes on the *x*-axis. Heatmap color indicates the number of superpopulations that share the same most frequent non-canonical motif. Cells without annotation or "." indicate that the most frequent pattern is a pure repeat of the canonical motif. Longer non-canonical motifs (>10 base pairs) are represented as "…". Uncolored cells marked with "." indicate allelic classes that are either not present (e.g., no UNK class at *FMR1*) or were not observed in the respective cohort (e.g., no premutations at *AFF2* in Cohort 1).

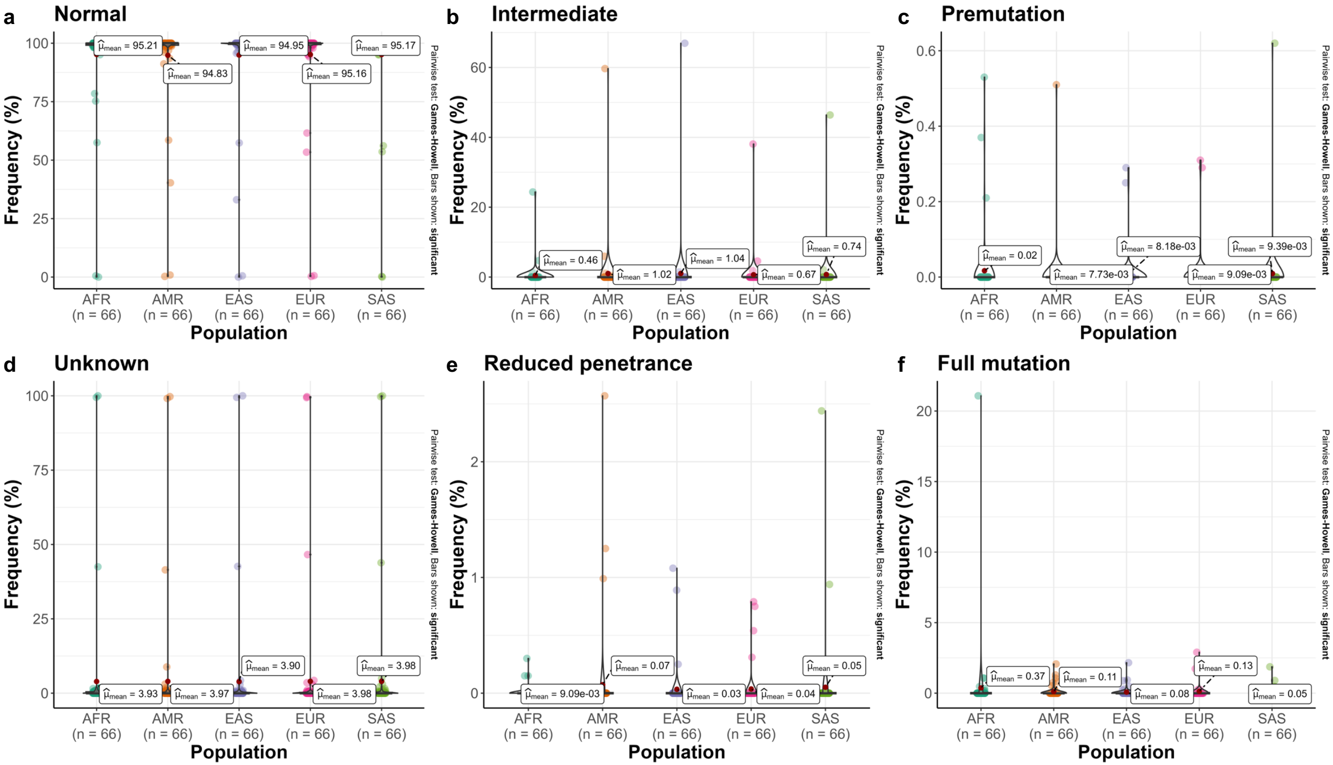


**Supplementary Fig. 3 | Allelic frequencies of disease-associated TRs (a–f).**

Annotated violin–boxplots with raw data overlay showing the frequencies of normal (**a**), intermediate (**b**), premutation (**c**), unknown (**d**), reduced penetrance (**e**), and full-mutation (**f**) alleles in Cohorts 1 and 2 across all loci. Comparisons of mean allelic frequencies (%) across populations are shown. Group differences were tested using one-way ANOVA, and pairwise comparisons were performed using the Games–Howell test with Bonferroni correction. The *n* values indicate the number of disease-associated TR loci studied.

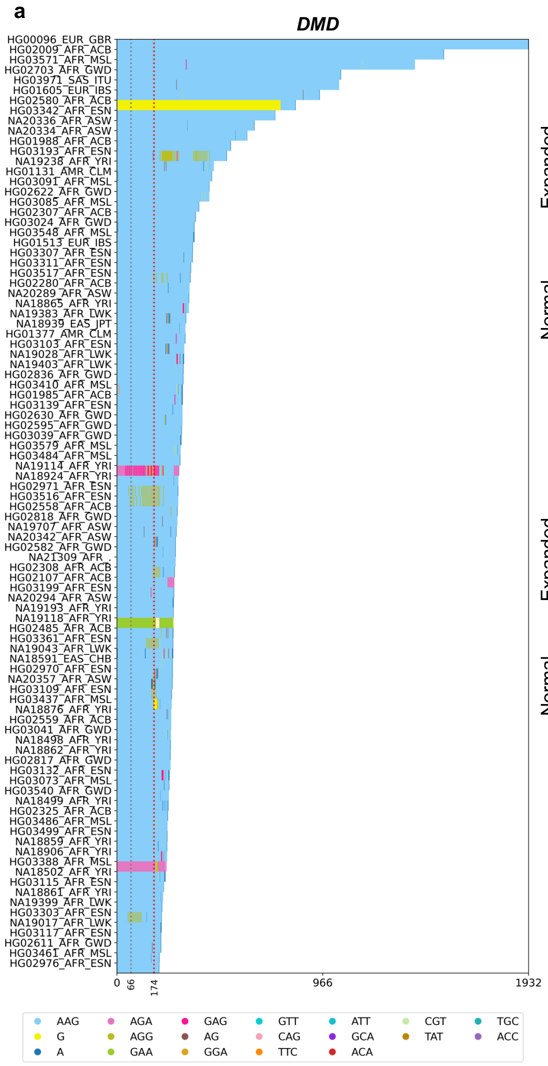


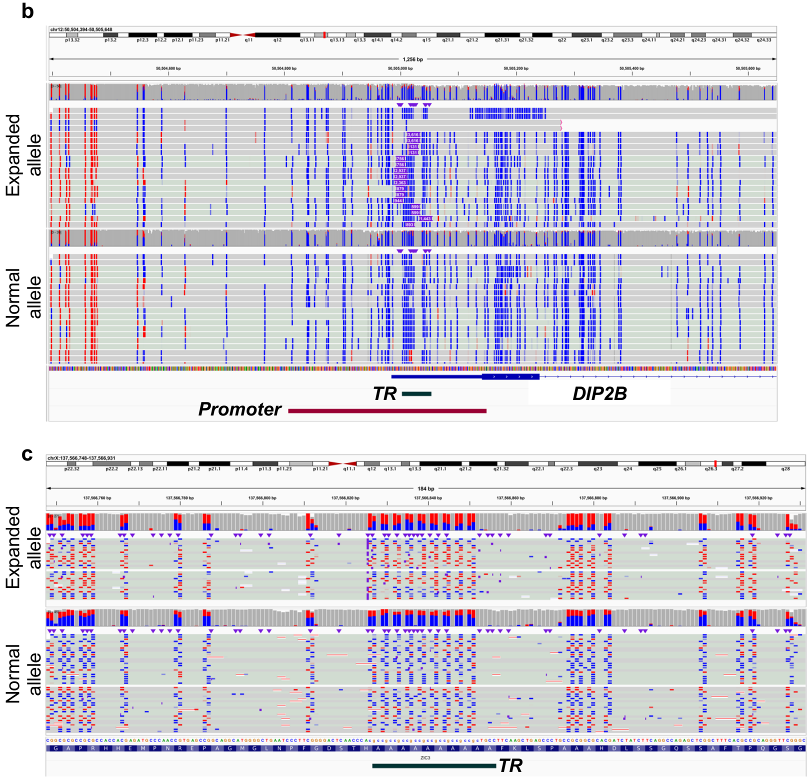


**Supplementary Fig.4 | Sequence composition and methylation profiles of expanded alleles at disease-associated TR loci (a–c).**

Sequence composition of expanded alleles at the *DMD* locus (**a**). Individual IDs, superpopulation, and population codes are indicated for each TR sequence (*y*-axis), sorted by repeat length (*x*-axis, in base pairs (bp)) with longer alleles at the top. The canonical motif is highlighted in light sky blue; each identified non-canonical motif is shown in a unique color. The bottom legend lists the top ten non-canonical motifs detected by TRMotifAnnotator. Vertical dotted gray and red lines indicate the upper bound of normal allele length and the lower bound of full-mutation allele length, respectively. Methylation profiles of normal and expanded *DIP2B* alleles in NA/GM20752 (**b**). IGV screenshot of the *DIP2B* promoter region showing methylation status around the expanded allele (top) and the normal haplotype (bottom). Multiple insertion signatures suggest repeat-length mosaicism, with observed allele sizes of ~600, 750, 900, 1500, and 3600 bp. Bases in red indicate methylated CpGs; blue bases indicate unmethylated CpGs. The $(CGG)_n$ TR and promoter positions are annotated below the panel. Methylation profiles of normal and expanded *ZIC3* alleles in HG00323 (**c**). IGV screenshot showing the expanded haplotype with 12 repeats (top) and the normal haplotype (bottom). Bases in red indicate methylated CpGs; blue bases indicate unmethylated CpGs. The TR position is annotated below the panel.

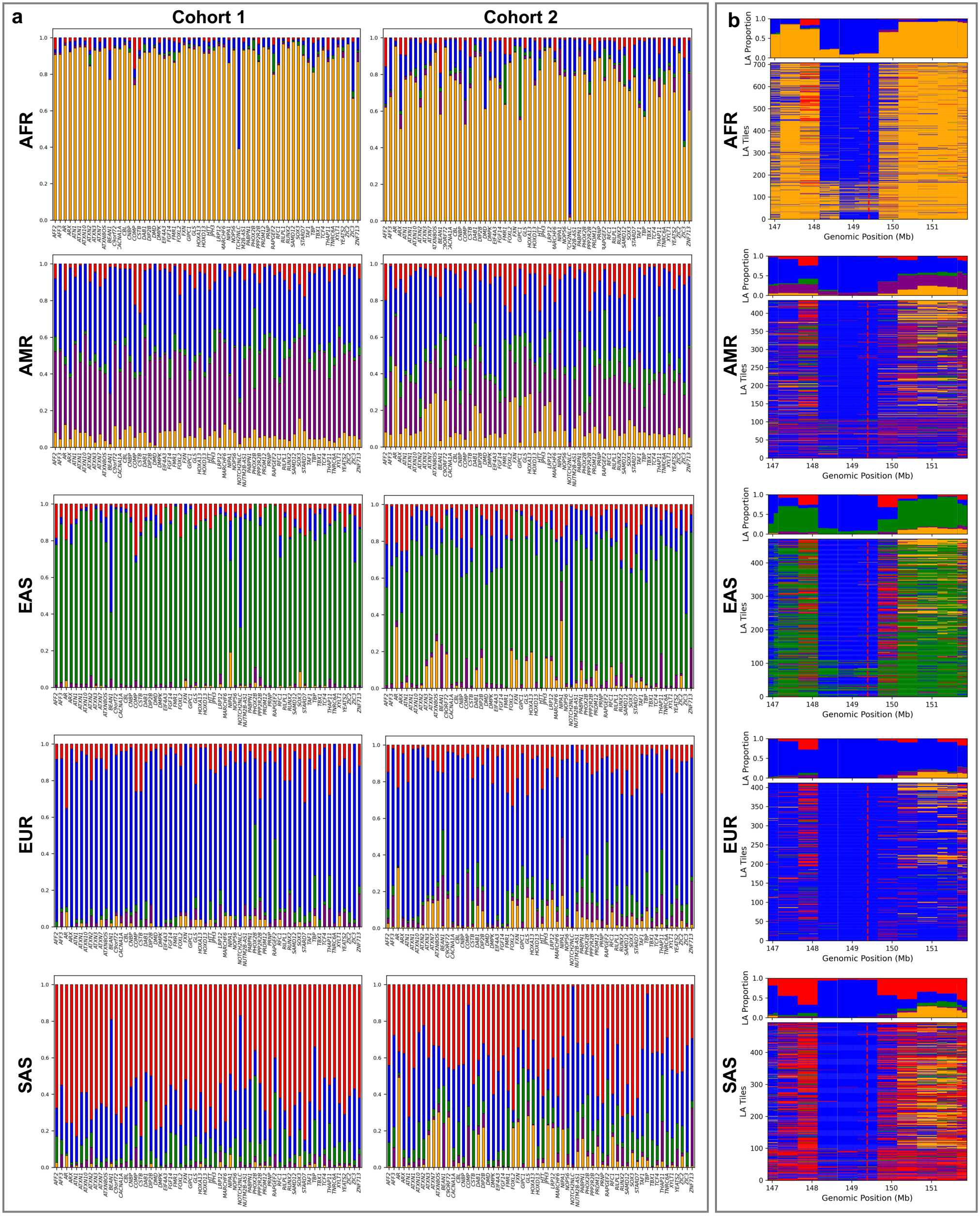


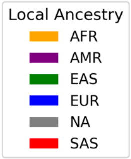


**Supplementary Fig. 5 | Local ancestries of disease-associated TRs (a,b).**

Local ancestry proportions within the 500 kb window encompassing each disease-associated TR locus, inferred by RFMix and stratified by superpopulation (**a**). Stacked bar plots show the proportions of AFR, AMR, EAS, EUR, SAS, or Unknown local ancestries (*y*-axis) at each TR locus (*x*-axis) in Cohort 1 (left) and Cohort 2 (right), with colors corresponding to local ancestry codes shown in the legend. Local ancestry in the 5 Mb region flanking the *NOTCH2NLC* TR, stratified by superpopulation (**b**). The top subpanel of each superpopulation shows stacked bar plots of local ancestry proportions (*y*-axis) across consecutive 500 kb windows within the 5 Mb region (*x*-axis). The bottom subpanel of each superpopulation shows a tile plot of local ancestry for each haplotype from Cohorts 1 and 2. The red dotted vertical line in the bottom panel.

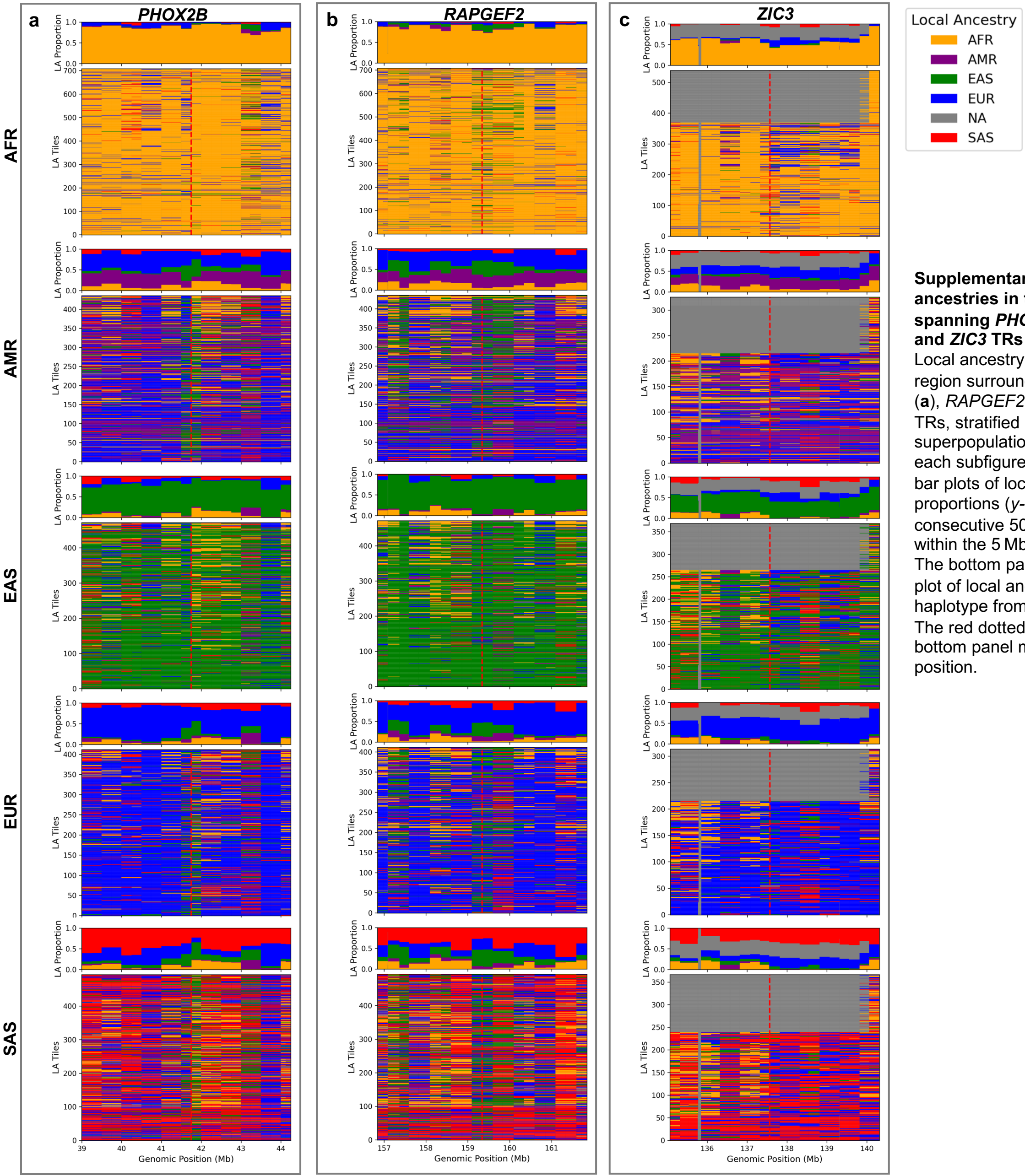


**Supplementary Fig. 6 | Local ancestries in the 5 Mb region spanning *PHOX2B*, *RAPGEF2*, and *ZIC3* TRs (a–c).**
Local ancestry across the 5 Mb region surrounding the *PHOX2B* (**a**), *RAPGEF2* (**b**), and *ZIC3* (**c**) TRs, stratified by superpopulation. The top panel in each subfigure shows stacked bar plots of local ancestry proportions (*y*-axis) across consecutive 500 kb windows within the 5 Mb region (*x*-axis). The bottom panel shows a tile plot of local ancestry for each haplotype from Cohorts 1 and 2. The red dotted vertical line in the bottom panel marks the TR start position.

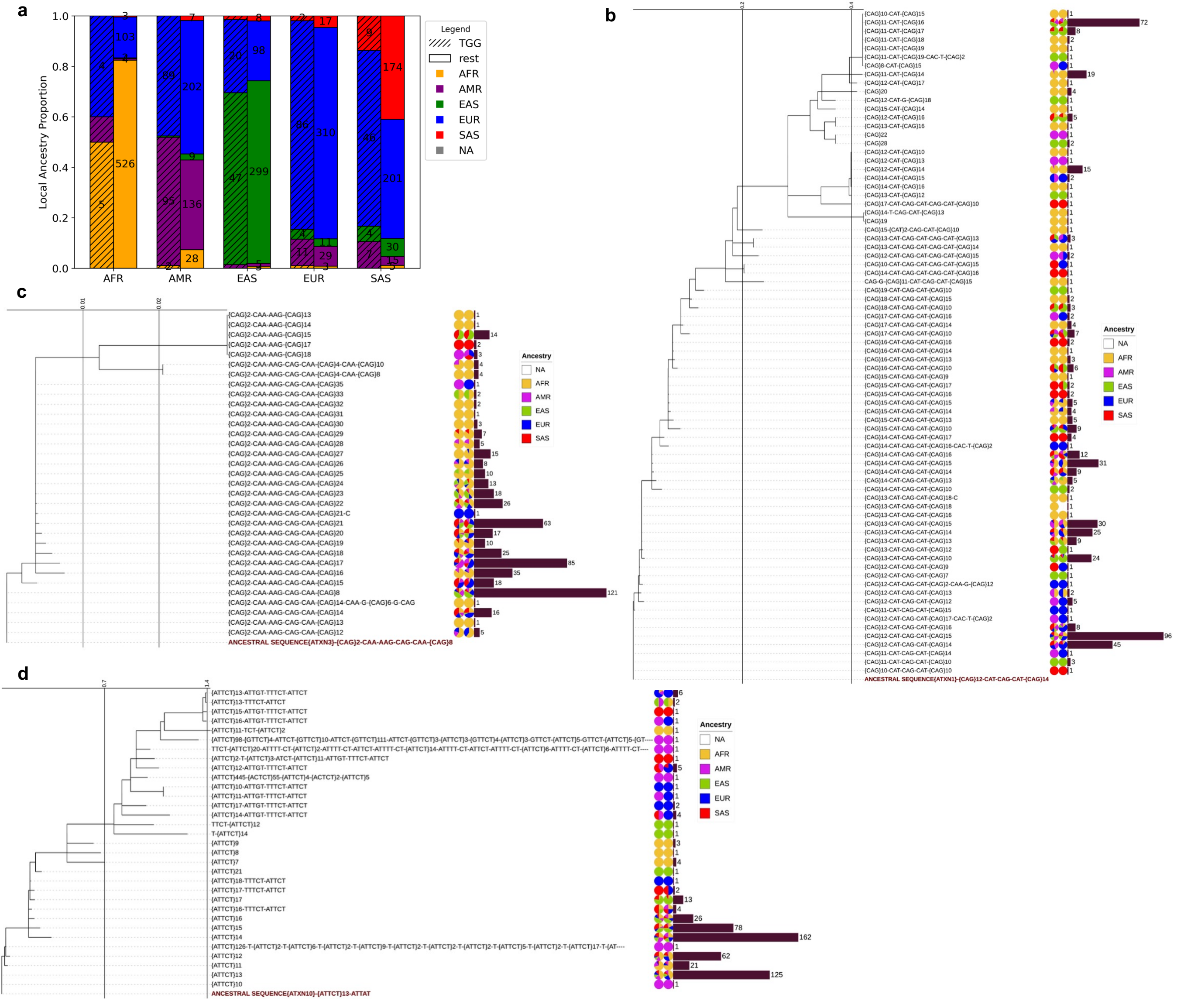


**Supplementary Fig. 7 | Inferred local ancestry of 16–17-repeat *TCF4* alleles and phylogenetic trees of *ATXN1*, *ATXN3*, and *ATXN10* repeat sequences (a–d).** Stacked bar plot showing local ancestry proportions of 16–17-repeat *TCF4* alleles with TGG interruptions, compared to all other alleles across superpopulations (**a**). Phylogenetic trees of *ATXN1* (**b**) *ATXN3* (**c**), and *ATXN10* (**d**) based on unique repeat structures and rooted with the ancestral sequence (bottom). Branch lengths are shown on the internal scale of each panel. Repeat structures are labeled at the tips. For each repeat structure, the first pie chart shows the proportion of alleles by superpopulation; the second pie chart shows the proportion of RFMix-inferred local ancestry. Adjacent bars indicate allele frequency by repeat structures. Colors for superpopulations and inferred local ancestries are defined in the legend and consistently applied to both.

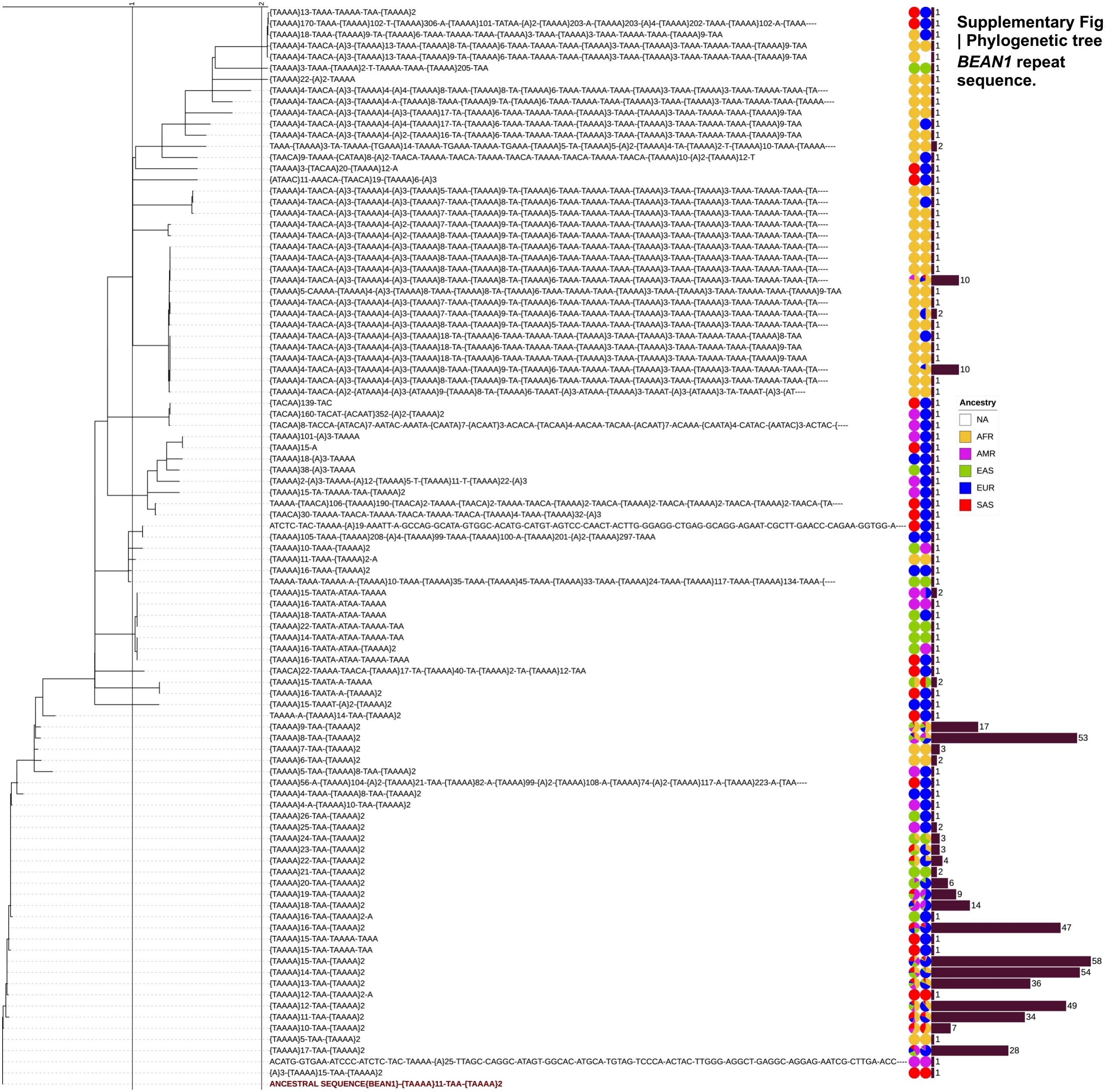


**Supplementary Fig. 8 | Phylogenetic tree of *BEAN1* repeat sequence.**

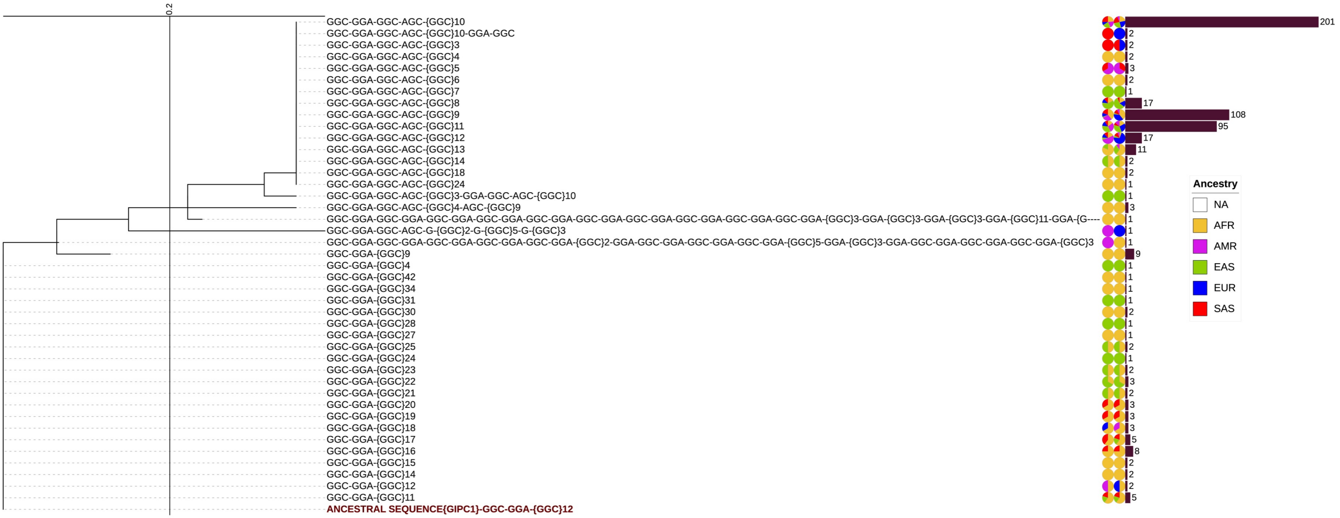


**Supplementary Fig. 9 | Phylogenetic tree of *GIPC1* repeat sequence.**

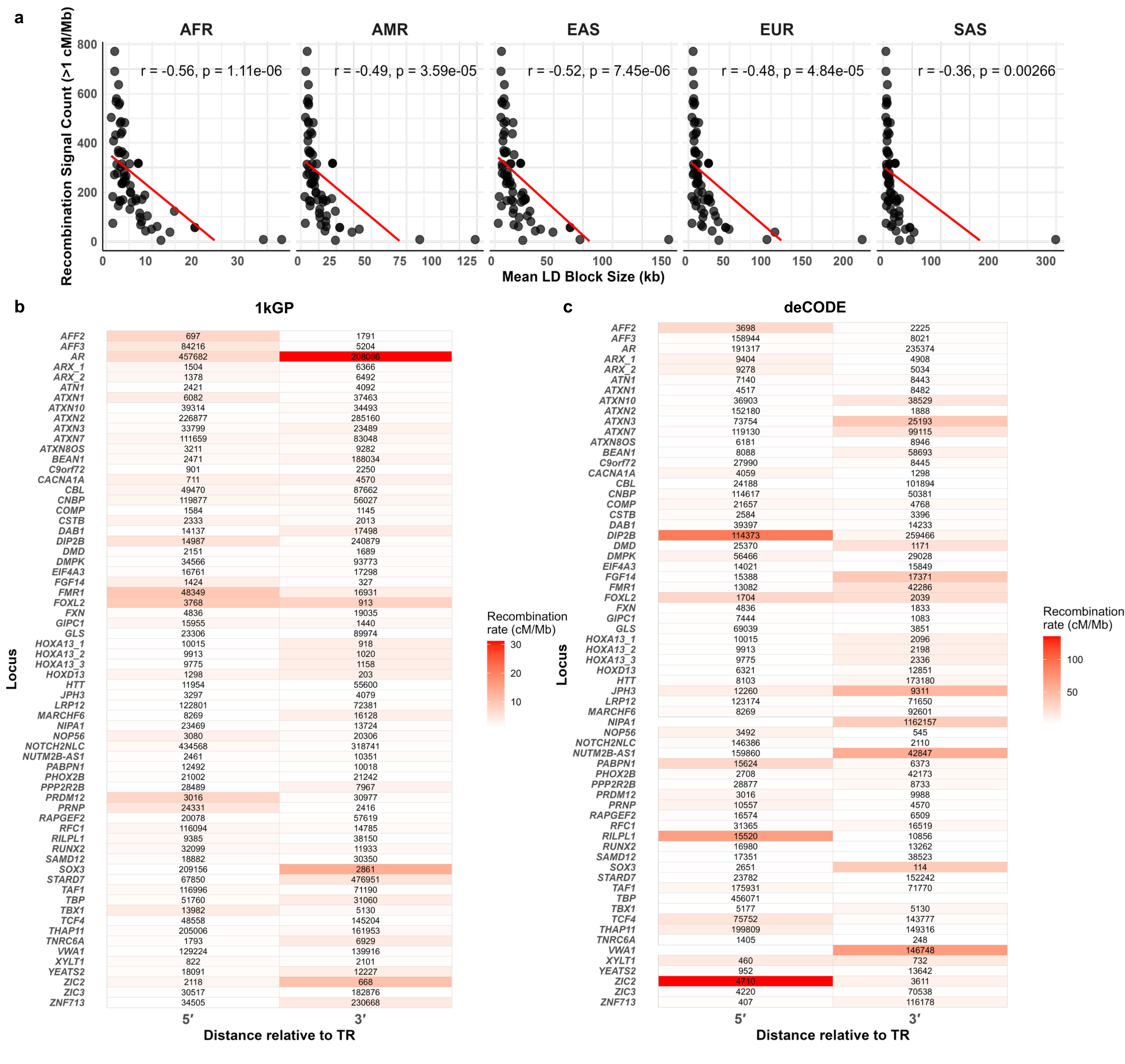
a
AFR
AMR
EAS
EUR
SAS
r = -0.56, p = 1.11e-06
r = -0.49, p = 3.59e-05
r = -0.52, p = 7.45e-06
r = -0.48, p = 4.84e-05
r = -0.36, p = 0.00266
Recombination Signal Count (>1 cM/Mb)
Mean LD Block Size (kb)
b
1kGP
Locus
AFF2 697 1791
AFF3 84216 5204
AR 457682 208066
ARX_1 1504 6366
ARX_2 1378 6492
ATN1 2421 4092
ATXN1 6082 37463
ATXN10 39314 34493
ATXN2 226877 285160
ATXN3 33799 23489
ATXN7 111659 83048
ATXN8OS 3211 9282
BEAN1 2471 188034
C9orf72 901 2250
CACNA1A 711 4570
CBL 49470 87662
CNBP 119877 56027
COMP 1584 1145
CSTB 2333 2013
DAB1 14137 17498
DIP2B 14987 240879
DMD 2151 1689
DMPK 34566 93773
EIF4A3 16761 17298
FGF14 1424 327
FMR1 48349 16931
FOXL2 3768 913
FXN 4836 19035
GIPC1 15955 1440
GLS 23306 89974
HOXA13_1 10015 918
HOXA13_2 9913 1020
HOXA13_3 9775 1158
HOXD13 1298 203
HTT 11954 55600
JPH3 3297 4079
LRP12 122801 72381
MARCHF6 8269 16128
NIPA1 23469 13724
NOP56 3080 20306
NOTCH2NLC 434568 318741
NUTM2B-AS1 2461 10351
PABPN1 12492 10018
PHOX2B 21002 21242
PPP2R2B 28489 7967
PRDM12 3016 30977
PRNP 24331 2416
RAPGEF2 20078 57619
RFC1 116094 14785
RILPL1 9385 38150
RUNX2 32099 11933
SAMD12 18882 30350
SOX3 209156 2861
STARD7 67850 476951
TAF1 116996 71190
TBP 51760 31060
TBX1 13982 5130
TCF4 48558 145204
THAP11 205006 161953
TNRC6A 1793 6929
VWA1 129224 139916
XYLT1 822 2101
YEATS2 18091 12227
ZIC2 2118 668
ZIC3 30517 182876
ZNF713 34505 230668
5′
3′
Distance relative to TR
Recombination rate (cM/Mb)
c
deCODE
Locus
AFF2 3698 2225
AFF3 158944 8021
AR 191317 235374
ARX_1 9404 4908
ARX_2 9278 5034
ATN1 7140 8443
ATXN1 4517 8482
ATXN10 36903 38529
ATXN2 152180 1888
ATXN3 73754 25193
ATXN7 119130 99115
ATXN8OS 6181 8946
BEAN1 8088 58693
C9orf72 27990 8445
CACNA1A 4059 1298
CBL 24188 101894
CNBP 114617 50381
COMP 21657 4768
CSTB 2584 3396
DAB1 39397 14233
DIP2B 114373 259466
DMD 25370 1171
DMPK 56466 29028
EIF4A3 14021 15849
FGF14 15388 17371
FMR1 13082 42286
FOXL2 1704 2039
FXN 4836 1833
GIPC1 7444 1083
GLS 69039 3851
HOXA13_1 10015 2096
HOXA13_2 9913 2198
HOXA13_3 9775 2336
HOXD13 6321 12851
HTT 8103 173180
JPH3 12260 9311
LRP12 123174 71650
MARCHF6 8269 92601
NIPA1 1162157
NOP56 3492 545
NOTCH2NLC 146386 2110
NUTM2B-AS1 159860 42847
PABPN1 15624 6373
PHOX2B 2708 42173
PPP2R2B 28877 8733
PRDM12 3016 9988
PRNP 10557 4570
RAPGEF2 16574 6509
RFC1 31365 16519
RILPL1 15520 10856
RUNX2 16980 13262
SAMD12 17351 38523
SOX3 2651 114
STARD7 23782 152242
TAF1 175931 71770
TBP 456071
TBX1 5177 5130
TCF4 75752 143777
THAP11 199809 149316
TNRC6A 1405 248
VWA1 146748
XYLT1 460 732
YEATS2 952 13642
ZIC2 4710 3611
ZIC3 4220 70538
ZNF713 407 116178
5′
3′
Distance relative to TR
Recombination rate (cM/Mb)

**Supplementary Fig. 10 | Recombination hotspots around disease-associated TR loci (a–c).**

Scatter plots depict the relationship between mean linkage disequilibrium block size (kb) and the number of recombination signals (>1 cM; 1kGP) within 1 Mb of each locus across five superpopulations (AFR, EUR, SAS, EAS, AMR) (**a**). Points represent loci, the red line shows the linear regression fit, and the Pearson correlation coefficient (r) and *p*-value are indicated in each panel. A similar trend was noted in deCODE (data not shown). Heatmaps showing distances from each TR midpoint to the nearest recombination hotspots (>1 cM/Mb) in 1kGP (**b**) and deCODE (**c**) datasets. Loci are listed on the *y*-axis; distances to the nearest 5’ and 3’ hotspots are shown for each TR. Heatmap cells display distances (in base pairs), with colors indicating recombination signal intensity as per the legend.

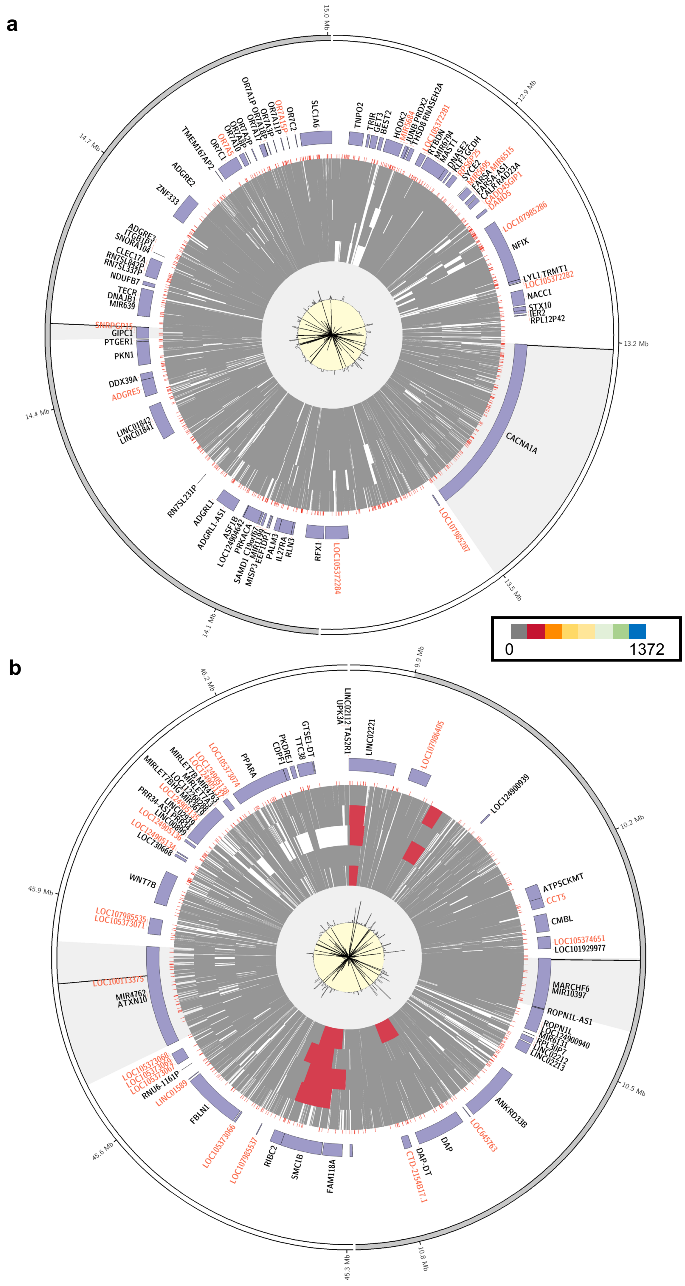


**Supplementary Fig. 11 | Linkage disequilibrium blocks around disease-associated TR loci (a,b).** Circos plots display, from outermost to innermost: the ideogram (Layer 1), gene track (Layer 2), and repeat track of simple repeats (Layer 3). Layers 4–8 show linkage disequilibrium blocks for each superpopulation: AFR, AMR, EAS, EUR, and SAS. Average recombination rates from 1kGP and deCODE Genetics (scale range: 1–100 cM/Mb) are shown in Layers 9 (light gray background) and 10 (light yellow background), respectively. All tracks are visualized across a 1-Mb region encompassing selected genes with disease-associated TRs. Gray highlights indicate the gene of interest, while black highlights within the gray mark the TR location. 1-Mb regions around loci shown are: *CACNA1A*, and *GIPC1* (**a**), and *ATXN10*, and *MARCHF6* (**b**). The color legend at the bottom corresponds to the number of SNVs (range: 0–1372) in the LD block heatmaps (Layers 4–8).

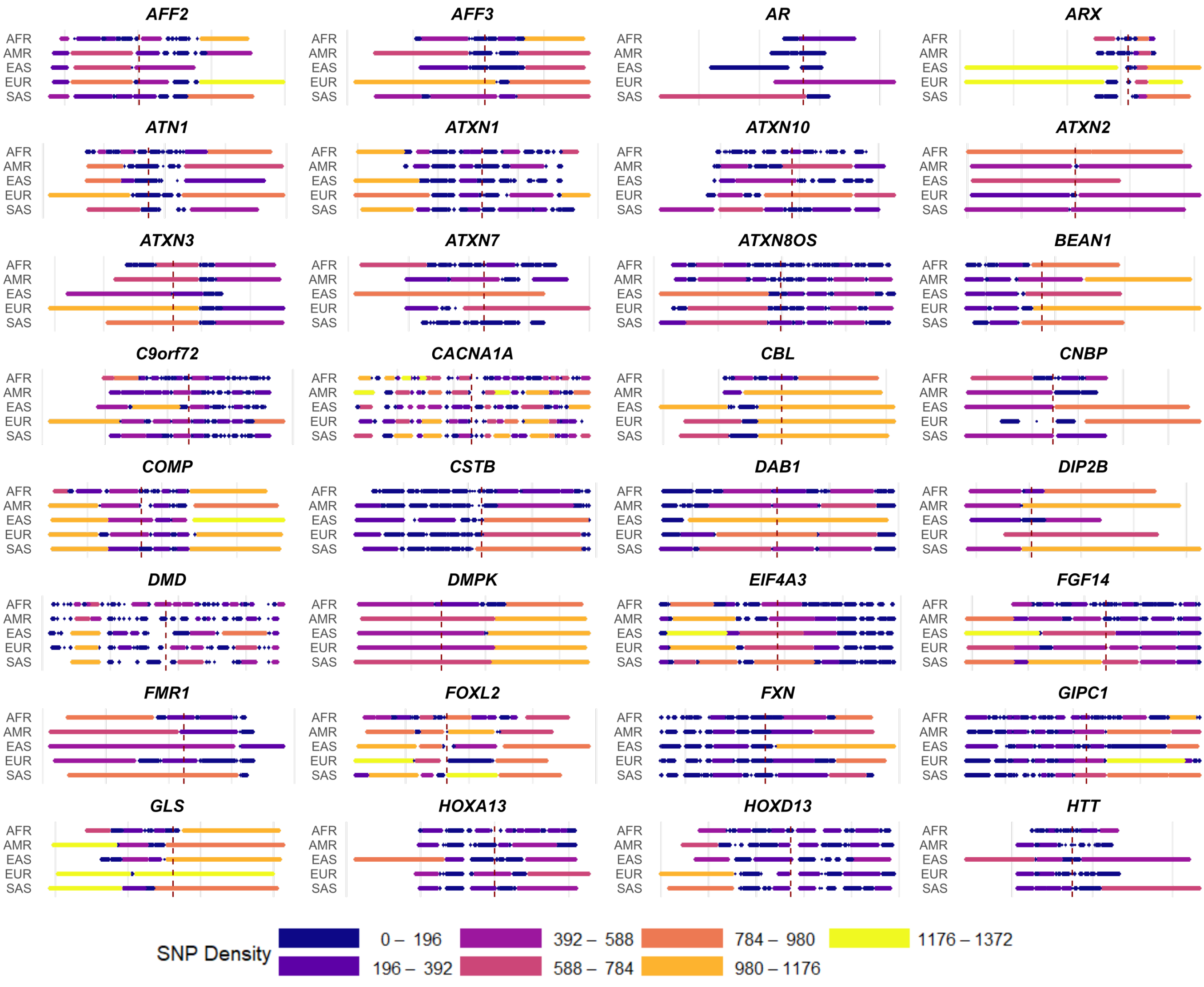


**Supplementary Fig. 12 | Linkage disequilibrium block patterns surrounding disease-associated TR loci.**
Each panel displays linkage disequilibrium block patterns and SNV density in the genomic regions flanking disease-associated TR loci across superpopulations (AFR, AMR, EAS, EUR, SAS). Horizontal colored segments show defined linkage disequilibrium blocks for each population, with color intensity indicating SNV density as shown in the accompanying color bar. *X*-axis shows ~100-kb window around each TR locus. The vertical dashed red line marks the exact position of the TR locus.

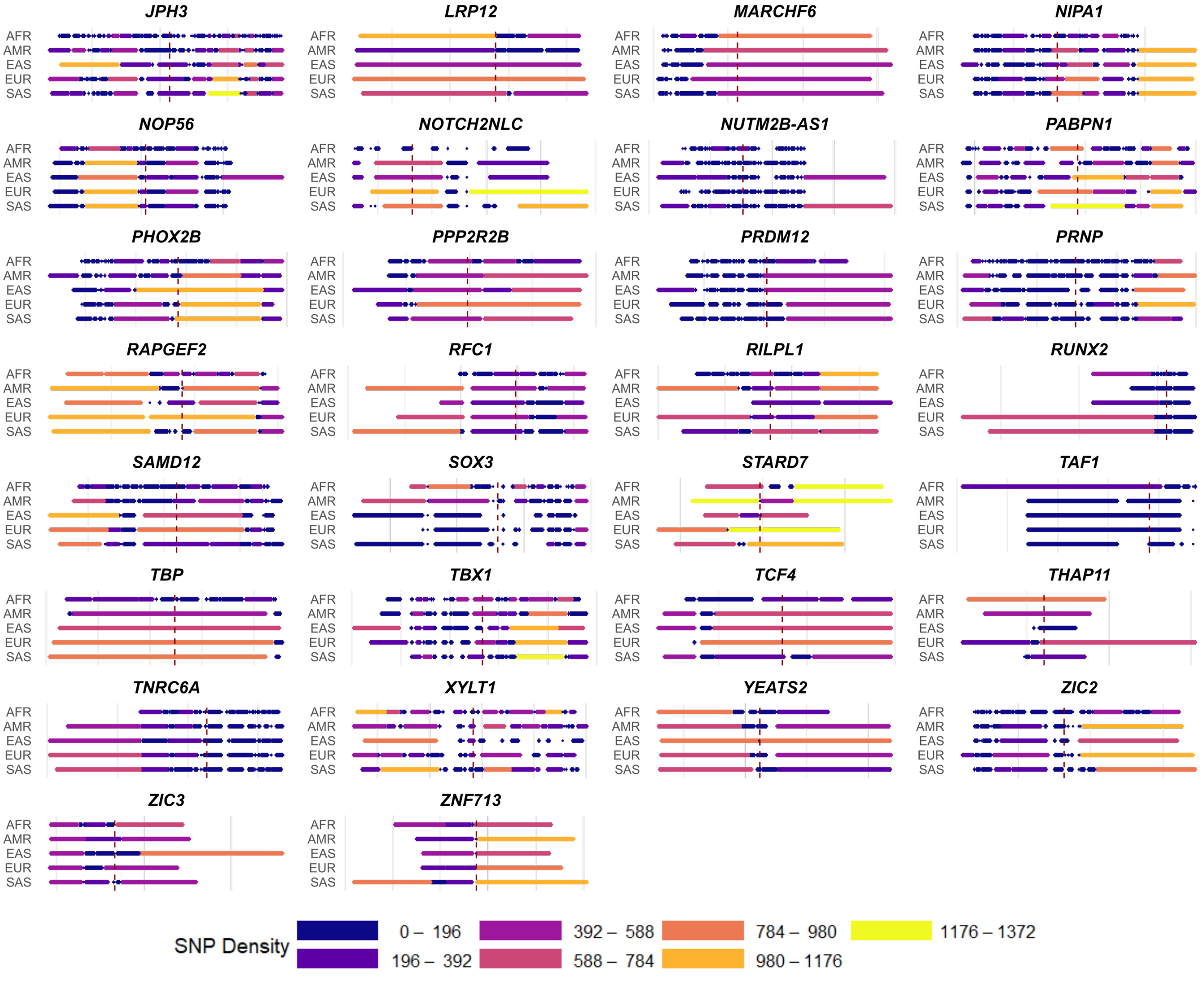


**Supplementary Fig. 13 | Linkage disequilibrium block patterns surrounding disease-associated TR loci (contd..).**
Panels show a continuation of Extended Data Fig. 9 with the remaining TR loci.

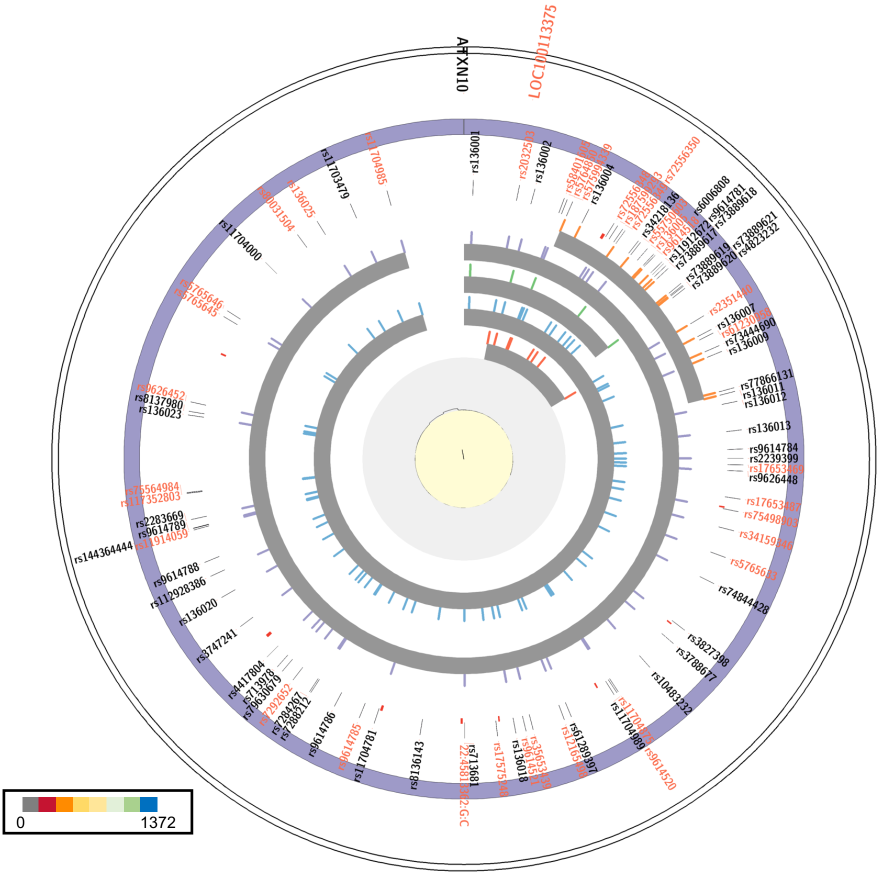


**Supplementary Fig. 14 | Linkage disequilibrium block patterns and SNV maps at the *ATXN10* TR locus.**
Circos plots depicting the genomic context of the *ATXN10* TR region on chromosome 22. The outermost layer (Layer 1) shows the chromosome ideogram, followed by the gene track (Layer 2), SNV track showing rsIDs or genomic coordinates of SNVs in linkage disequilibrium block (Layer 3), and repeat annotation track for simple repeats (Layer 4). Layers 5–14 show the SNVs within linkage disequilibrium block and linkage disequilibrium block structure from PLINK for each superpopulation (AFR, AMR, EAS, EUR, SAS). Average recombination rates from 1kGP and deCODE Genetics (scale range: 1–100 cM/Mb) are shown in the innermost layers in light gray and yellow backgrounds, respectively. The color scale at the bottom indicates the number of SNVs (ranging from 0 to 1372) in linkage disequilibrium block heatmaps.
